# CCDC78: unveiling the function of a novel gene associated to hereditary myopathy

**DOI:** 10.1101/2023.12.23.23300356

**Authors:** Diego Lopergolo, Gian Nicola Gallus, Giuseppe Pieraccini, Francesca Boscaro, Gianna Berti, Giovanni Serni, Nila Volpi, Patrizia Formichi, Silvia Bianchi, Denise Cassandrini, Vincenzo Sorrentino, Daniela Rossi, Filippo Maria Santorelli, Nicola De Stefano, Alessandro Malandrini

## Abstract

*CCDC78* was indicated about ten years ago as novel candidate gene for the autosomal dominant centronuclear myopathy-4 (CNM4). However, to date, only one family has been described and CCDC78 function remains unclear. Here we deeply analyze for the first time a family harbouring a *CCDC78* nonsense mutation. Histopathological features included, as novel histological hallmark, peculiar sarcoplasmic reticulum (SR) abnormalities. We provided evidence of nonsense mediated mRNA decay, defined novel *CCDC78* transcripts and, through transcriptome profiling, detected 1035 muscular differentially expressed genes including a series of genes involved in SR. Through coimmunoprecipitation assay and mass spectrometry studies we demonstrated that CCDC78 interacts with two pivotal SR proteins: SERCA1 and CASQ1. We also found an interaction with MYH1, ACTN2 and ACTA1. Our findings shed light on interactors and possible role of CCDC78 in skeletal muscle, thus allowing us to locate the protein in SR and to consider *CCDC78* as CNM4 causative gene.

## Introduction

Congenital inherited myopathies are a large and heterogeneous group of muscle disease affecting the skeletal muscle tissue^1–3^. They arose by genetically determined muscle protein defects and classified based on muscle biopsy findings^2^. In the last decades advances in molecular genetics have allowed to identify an increasing number of causative genes linked to different types of hereditary myopathies^4^ and it has become evident that mutations in a same gene can lead to more than one pathological and clinical phenotype as well as the same pathological feature can result from mutations in different genes.

The coiled-coil domain-containing protein 78 gene (*CCDC78)*, mapped by Daniels et al.^5^ to chromosome 16p13.3, encodes a protein previously reported as involved in centriole amplification in multiciliated cells^6, 7^ and in skeletal muscle function ^8^. Majczenko et al.^8^ found a predominant gene expression in human skeletal muscle and they showed by immunostaining a reticular pattern partially overlapping the triad, the intersection of the T-tubule and the terminal sarcoplasmic reticulum (SR). However, only one *CCDC78* mutated family has been reported to date^8^: the patients, harboring a splice-acceptor variant, showed early-onset distal muscle weakness, myalgia, and easy fatigue with normal serum creatine phosphokinase (CPK) levels. Muscle biopsy revealed CCDC78 accumulation in large sarcoplasmic aggregates, which costained for desmin, actin, and strongly for RyR1; CCDC78 localized to the sarcolemmal membrane and the perinuclear region, as well as in a latticework-type pattern within the sarcoplasm. Costaining with RyR1 reveals that the latticework pattern corresponded to the triad, the intersection of the terminal SR and the T-tubule. The authors thus suggested a possible interplay between RyR1 and CCDC78. Based on these data, *CCDC78* was defined as a new candidate gene for the autosomal dominant centronuclear myopathy-4 (CNM4; OMIM#614807). However, to date, the role of *CCDC78* in muscle function remains unclear; furthermore, the possible interaction of CCDC78 with RyR1 remains uncertain and yet to be demonstrated.

Here we deeply analyzed a family harbouring the first nonsense mutation in *CCDC78*. We performed optic and electron microscopy study and multiple immunofluorescence assays on muscle biopsy. We analyzed *CCDC78* transcripts and protein in different tissues from healthy and mutated subjects, and we performed transcriptome profiling by RNA-seq and RT-qPCR from muscle tissues. We explored whether such nonsense mutation acts with a loss of function or a gain-of-function mechanism and the possible activation of a non-mediated decay (NMD) mechanism. Lastly, after co-immunoprecipitation (Co-Ip) assay and mass spectrometry analysis we defined the major interacting proteins of CCDC78.

## Results

### 1. The nonsense *CCDC78* mutation leads to a dystrophic muscle process mainly involving the SR

Our patient, a male in his 50s, referred from the age of about 40-45 years the presence of myalgias, continuous painful muscle cramps in the calf and foot muscles, excessive sweating, and easy fatigue. Blood chemistry showed persistent CPK elevation: the last two CPK measurements were 655 and 663 UI/l, respectively; myoglobin was 96 ng/ml. Aspartate aminotransferase and alanine aminotransferase were 36 IU/l and 56 IU/l, respectively. A liver ultrasound revealed mild hepatic steatosis. The neurological examination showed lively deep tendon reflexes in the lower limbs and slight hypertrophy of the calves bilaterally (fig. 1b-d) without strength deficit. The EMG study was normal. A patient’s relative was reported to have had senile dementia, epilepsy and bilateral calf hypertrophy. Another patient’s relative had a patent foramen ovale and bilateral calf hypertrophy (fig. 1a).

**Fig. 1.**
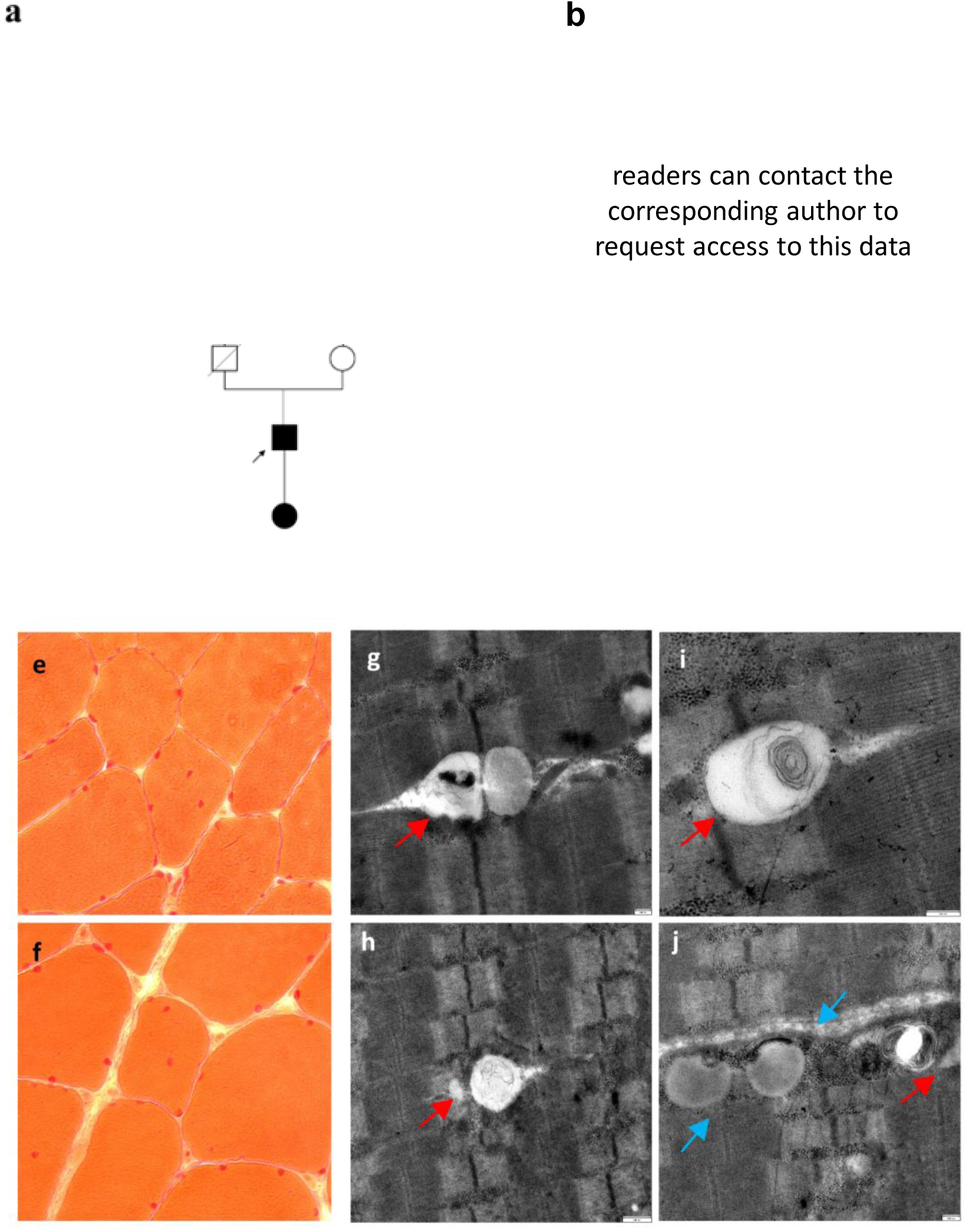
Pedigree of the CCDC78 mutated patients **(a)**, photographs of the proband showing slight hypertrophy of the calves bilaterally **(b-d)**, muscle biopsy with E-H staining (20X) showing nuclear centralizations **(e-f)** and TEM (scale bar= 200 nm) showing peculiar dilated terminal SR (red arrows), whirls of redundant membranes (green arrow) and areas of dilated and swollen SR with numerous areas of abnormal accumulations of membranous material (blue arrows).

The muscle biopsy from left vastus lateralis muscle showed inhomogeneity of caliber and shape of the muscle fibers, with hypotrophic and hypertrophic fibers. Neither cellular necrosis nor inflammatory infiltrates were observed. Nuclear centralizations were present in 6-7% of muscle fibers (fig. 1e-f). Gomori’s trichrome stain and oxidative reactions were normal. No increase in PAS positive or sudanophilic material was detected. A predominance of type 2 fibers was evident. The immunohistochemical study with dystrophin 2, dystrophin 3, caveolin, dysferlin, 35 kDa delta-sarcoglycan, 35 kDa gamma-sarcoglycan, 43 kDa beta-sarcoglycan and 50 kDa alpha-sarcoglycan antibodies was normal. Transmission electron microscopy (TEM) revealed peculiar, dilated terminal SR, whirls of redundant membranes and areas of dilated and swollen SR with numerous areas of abnormal accumulations of membranous material (fig. 1g-j). Immunofluorescent analysis on muscle tissue showed CCDC78 aggregates, overlapping with RyR1 (fig. 2) and desmin.

**Fig. 2.**
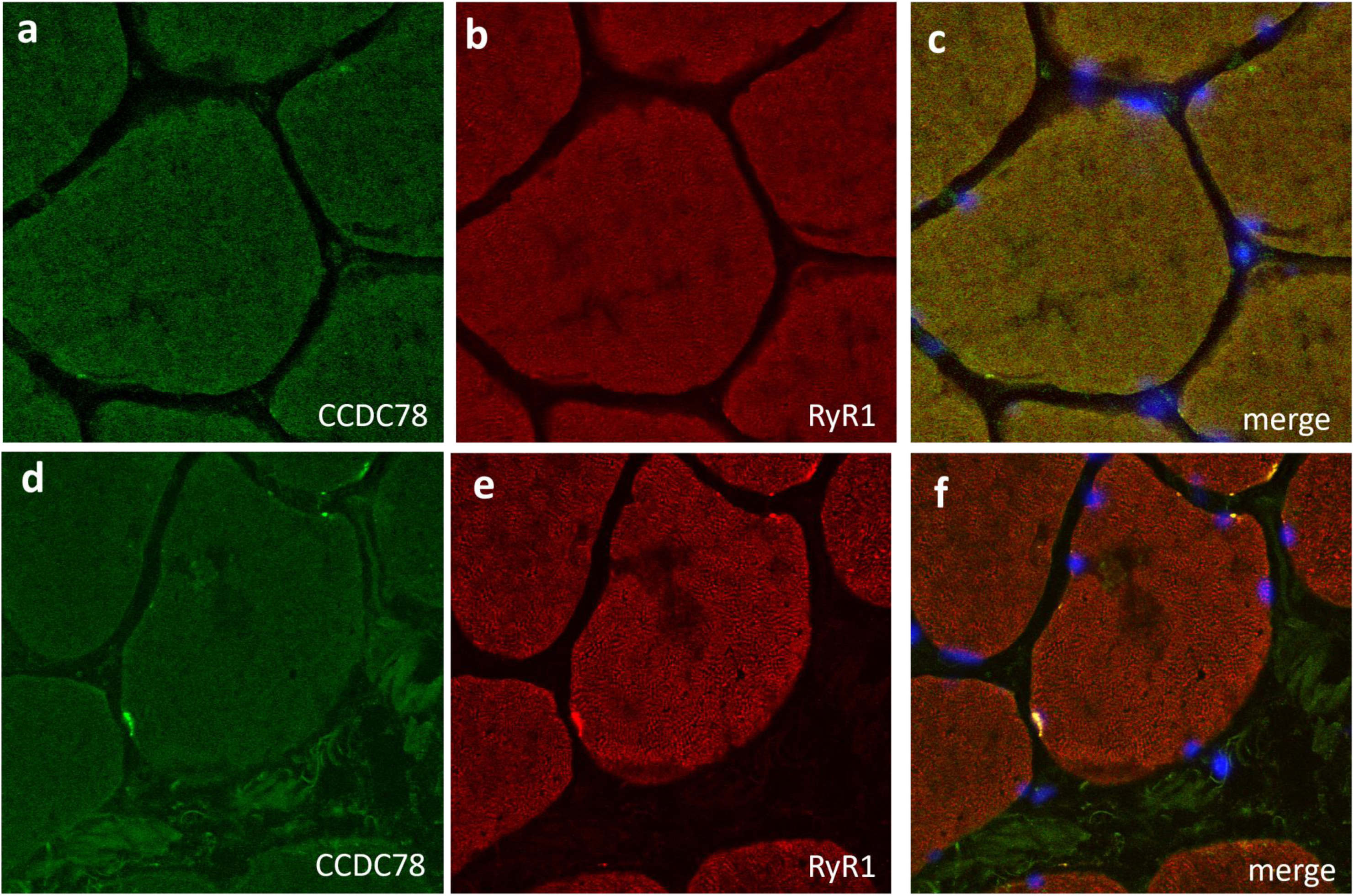
Immunofluorescent analysis on muscle tissue (20X). By comparing control **(a-c)** and CCDC78 mutated muscles **(d-f)** after CCDC78 **(a,d)** and RyR1 staining **(b,e)**, we showed CCDC78 aggregates, overlapping with RyR1.

The WES revealed in our patient the heterozygous mutation c.1206G>A (p.Trp402*) located in exon 13 of *CCDC78* gene (rs752371476; NM_001031737.3) resulting in a premature stop codon at position 402 out of 438.

Two alternative *CCDC78* transcripts are actually known: NM_001378030.1 and NM_001031737.3. The NM_001378030.1 transcript (fig. 3a, in yellow), having 1611 bp length, codes for the 438 aa protein isoform; the NM_001031737.3 transcript (fig. 3a, in blue) has a 1585 bp length and, differently from the previous one, includes four nucleotides (GCAG) at the beginning of exon 13 and codes for the 470 aa protein isoform.

**Fig. 3.**
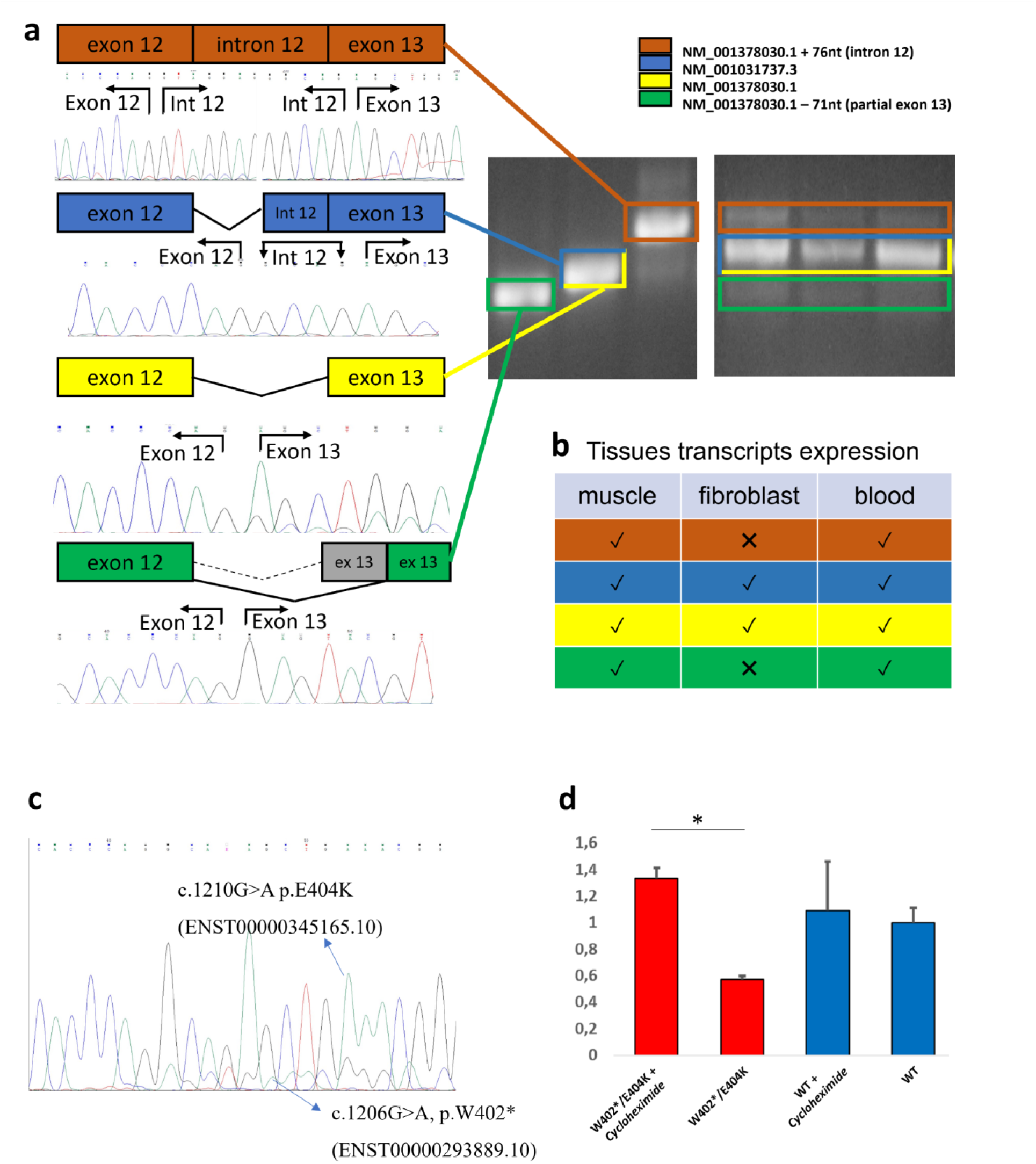
PCR amplification of CCDC78 exons 10-14 showed the presence of four transcripts **(a)**. Agarose gel electrophoresis (2% agarose) of PCR amplified products using specific PCR primer sets. Lanes 1-3 showed the sequenced PCR products in muscle; lanes 4, 5 and 6 showed transcript pattern of exons region 10-14 in PBLs, muscle and fibroblasts, respectively. Tissues trascripts in muscle, fibroblast and blood are shown (v = transcript sequenced, X = transcript not sequenced **(b)**. In order to assess NMD due to p.W402* mutation, suggested by an apparently lower level of transcript NM_001031737.3 from muscle sample **(c)**, we analyzed the CCDC78 expression in lymphocytes of the CCDC78 mutated patient and control subjects in basal conditions and after cycloheximide treatment. The patient showed a significant increase in transcript values compared to the control **(d)**. Relative expression levels were calculated relative to HPRT1, SERPINC1 and ZNF80 mRNA levels and set to 1. (n=3, p<0.01).

The identified *CCDC78* variant was predicted to be damaging by CADD-phred prediction tools (CADD-phred=44) and BayesDel algoritm (addAF score 0.5116, noAF score 0.5125). The Genomic Evolutionary Rate Profiling (GERP) score was 1,96 thus indicating that the variant may be under evolutionary constraint. The minor allele frequency (MAF) was <0.01%, the ExAc all frequency of the variant was 0.00001671%, gnomAD all was 8.01218e-06. Notably, the mutation results as a missense variant in the longer isoform of the transcript: c.1210G>A (p.Glu404Lys, NM_001378030.1). The variant was Sanger confirmed. Variant segregation analysis showed the mutation only in the young relative.

### 2. Human tissues express several *CCDC78* isoforms and the nonsense *CCDC78* variant results in a mutant allele mostly degraded by NMD

Several PCR including exons 10-14 were set up to study the 3’ region of the transcript, where the mutation is located. PCR were performed on RNA extracted from fibroblasts, lymphocytes, and muscle tissues deriving from the patient and controls. We identified both NM_001031737.3 and NM_001378030.1 transcripts. Moreover, we also detected two further in-frame transcripts due to alternative splicing involving the last exons of the gene: the first transcript derived from retention of intron 12 (NM_001378030.1:r.1201_1275del), the second transcript derived from activation of a new acceptor site in exon 13 (NM_001378030.1:r.1201_1275del) (fig. 3a-b).

To assess if the stop codon leads to NMD, as suggested by an apparently lower level of NM_001031737.3 transcript, we analyzed the *CCDC78* expression in the lymphocytes of the patient and a control subject comparing the gene expression at basal conditions and after treatment with cycloheximide, a known NMD inhibitor. The patient showed a significant increase in transcript values after treatment compared to the control (n=3, p<0.01) (fig.3c-d) thus indicating the activation of NMD.

CCDC78 expression analysis by WB in the *CCDC78* mutated patient and three controls showed the presence of different protein isoforms at 52 (Uniprot: H3BLT8), 48 (Uniprot: A2IDD5-1) and 37 kDa (Uniprot: A2IDD5-5) (fig. 4a). Only the 52 kDa isoform in the *CCDC78* mutated patient was significantly reduced compared to controls (36.85±11.61%, p-value <0.01). The 48 and 37 CCDC78 isoforms were respectively 84.33±2.75% (SD) and 114.28±2.75% (SD) compared to controls (fig. 4b).

**Fig. 4.**
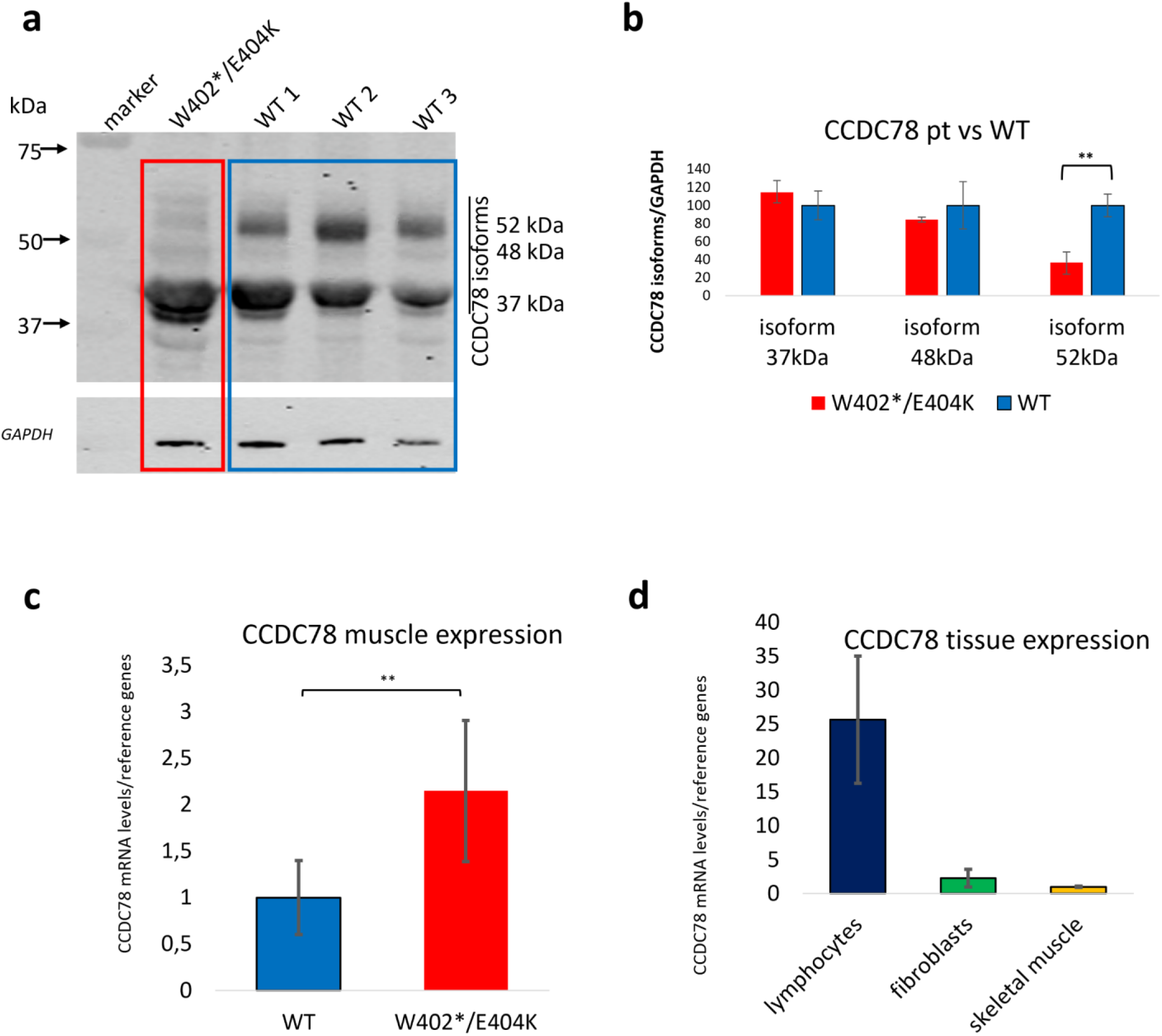
Western blot (WB) analysis showing the expression levels of CCDC78 isoforms in muscle tissue of patient and three controls **(a).** The bar graph **(b)** shows the isoforms expression fold change of CCDC78, calculated by setting the ratio of CCDC78 protein/GAPDH protein band intensities in the control group to 100. The bars show the mean ± SD. (n≥5, *p < 0.01). RT-qPCR in muscle showed a significant increase in our patient **(c)** calculated by setting the ratio of CCDC78/reference genes expression in the control group to 1. Relative expression levels were calculated relative to *HPRT1*, *SERPINC1* and *ZNF80* mRNA levels. The bars show the mean ± SD (n=2, **p < 0.01). RT-qPCR showed the relative expression levels of CCDC78 in lymphocytes, fibroblasts, and muscle of control samples **(d)**. CCDC78 expression in muscle tissue was set to 1 (SD ± 0.09), the relative expressions in fibroblasts and lymphocytes were respectively 2.3 (SD ± 1.30045) and 25.62 (SD ± 9.39). Relative expression levels were calculated relative to *HPRT1* and *ZNF80* mRNA levels (n=2).

RNA extracted from human fibroblasts, lymphocytes and muscle tissues were analyzed via RT[qPCR to evaluate the amount of *CCDC78* gene expression. RT-qPCR in muscle showed a significant expression increase in our patient compared to healthy controls (p<0.01) (fig. 4c). Moreover, the analysis of relative expression of *CCDC78* in lymphocytes, fibroblasts, and muscle tissue from healthy controls revealed a lower *CCDC78* expression in muscle tissue. Indeed, by setting muscle tissue expression to 1 (SD ± 0.09), the relative expressions in fibroblasts and lymphocytes were 2.30 (SD ± 1.30) and 25.62 (SD ± 9.38), respectively (fig. 4d). Accordingly, *CCDC78* resulted to be very low expressed in muscle tissue by the RNA-seq analysis we performed. These findings are in line with previous data of Gonorazky et al^9^ that showed in GTEx skeletal-muscle controls and in a large cohort of muscle biopsies 15 genes including *CCDC78* expressed at <1 RPKM.

Through Dense Alignment Surface (DAS) method^10^ we predicted possible CCDC78 transmembrane segments (TS). For the wild type (WT) 48 kDa, WT 52 kDa and mutated 52 kDa isoforms the potential TS lies between position 209 and 216 (1.7 cutoff). Only the mutated 48 kDa isoform showed a potential TS between position 210 and 216 (1.7 cutoff) (fig. 5). To define possible structural effect of the variants, 3D models of wild-type human CCDC78 (both NM_001031737.3 and NM_001378030.1) and its variants (p.Trp402*, NM_001031737.3 and p.Glu404Lys, NM_001378030.1) were generated through SwissModel (https://swissmodel.expasy.org/). Our data showed a critical change in 3D structure driven by both p.Trp402* (NM_001031737.3) and p.Glu404Lys (NM_001378030.1). Other than a reduced protein length for p.Trp402*, as expected, we observed for both the variants a different spatial orientation of the first (Glu56-Asp105) and the fourth (Asp381-Ser401) alpha-helixes respect to the second one (Asn156-Asp259), likely to contain the predicted TS (fig. 5). This effect may lead to a dysfunctional CCDC78 protein. Moreover, the analysis of the mutated 52 kDa isoform by HOPE (https://www3.cmbi.umcn.nl/hope/) indicated that the Lys404 aminoacidic residue, bigger than the wild-type Glu404 residue, might lead to bumps on the 3D structure of the protein near a highly conserved position of CCDC78, thus suggesting a possibly damaging effect. Moreover, the Glu>Lys substitution, swapping a negative to positive residue, may further affect protein function trough repulsion of other residues with the same charge.

**Fig. 5.**
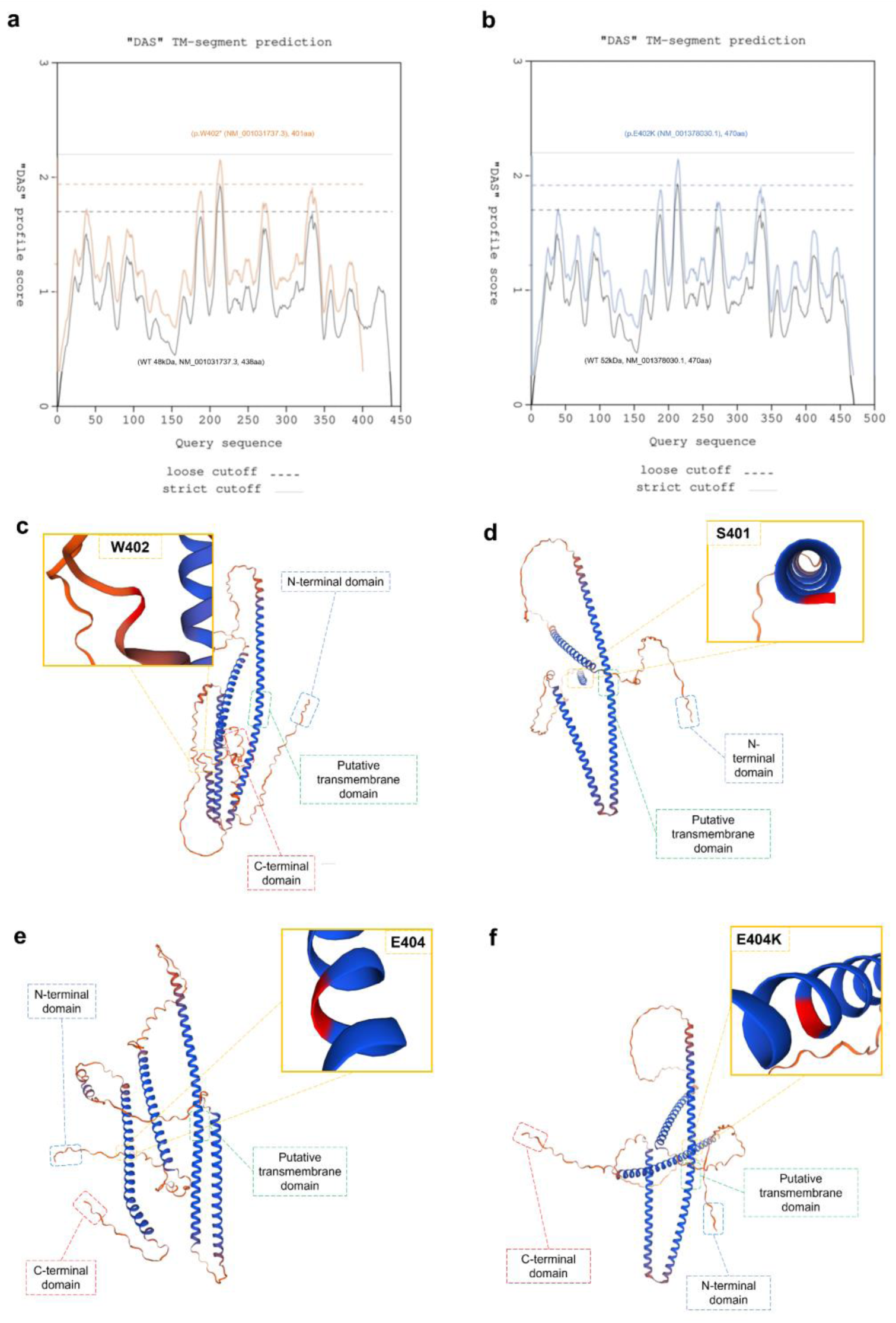
Curves obtained by pairwise comparison of the proteins in the test set in “each against the rest” fashion (**a,b**): there are two cutoffs indicated on the plots: a “strict” one at 2.2 DAS score, and a “loose” one at 1.7. The hit at 2.2 is informative in terms of the number of matching segments, while a hit at 1.7 gives the actual location of the transmembrane segment (TS). The segments reported in the feature table (FT) records of the SwissProt database are marked at 1.0 DAS score (”FT lines”). In **(a)** we reported a staggered superposition of the potential TS predictions for both the WT 48kDa protein (NM_001031737.3) (black lines) and the relative mutated protein (p.Trp402* (NM_001031737.3)) (orange lines): WT TS starts at 209 and stops to 216 (length ∼8, cutoff ∼ 1.7); in the mutated protein TS starts at 210 and stops to 216 (length ∼7, cutoff ∼ 1.7). The plots are very similar; however, the TS is shorter in the mutated protein. In **(b)** we reported a staggered superposition of the potential TS predictions for both the WT 52kDa protein (NM_001378030.1) (black lines) and the relative mutated protein (p.Glu404Lys (NM_001378030.1)) (blue lines): both WT and mutated TS start at 209 and stops to 216 (length ∼8, cutoff ∼ 1.7). The plots are identical and perfectly overlapping. **3D modelling**: both the WT isoforms and the relative mutated protein are shown (A2IDD5_CCDC78_HUMAN, 438aa, WT **(c)**, A2IDD5_CCDC78_HUMAN, 401aa, p.Trp402* **(d)**, H3BLT8_CCDC78_HUMAN, 470aa, WT. **(e)**, H3BLT8_CCDC78_HUMAN, 470aa, p.Glu404Lys **(f)**. For both the variants we observed a different spatial orientation of the first (Glu56-Asp105) and the fourth (Asp381-Ser401) alpha-helixes respect to the second one (Asn156-Asp259).

### 3. The *CCDC78* mutation induces upregulation of genes involved in SR

We explored the transcriptome of the *CCDC78* mutated patient by comprehensive RNA-seq analyses using muscle tissue from the patient and three controls to assess differential gene expression changes associated with *CCDC78* mutation. The box plot analysis shows a visual representation of raw (fig. 6a) and normalized (fig. 6b) expression values. The figure 6c shows the distances measured using expression values from each sample: according to this sample distance method, the shorter the distance, the more closely related the samples are. This method allowed us to identify if the two groups are closely related or not: Euclidean distance analysis followed by hierarchical clustering analysis indicated that each set of replicates clustered together. Principal component analysis (PCA) was performed on the covariance matrix of 14’858 muscular expressed genes in our four samples. PCA reveals gene expression differences among the two experimental conditions confirming that control replicates well clustered together and that gene expression was mainly separated by health condition (PC1, 60% variance) (fig. 6d).

**Fig. 6.**
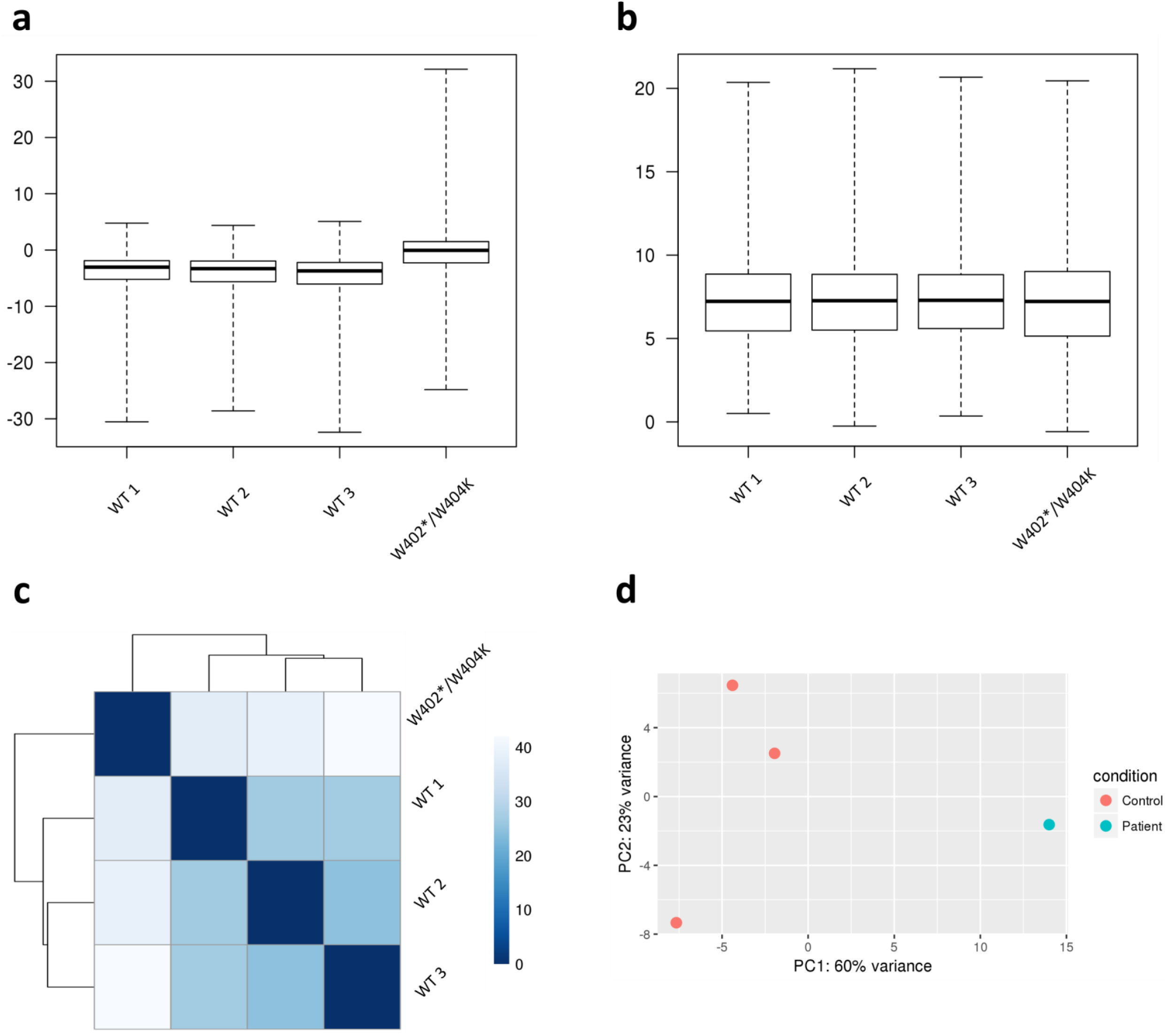
Comparison of gene expression between the defined groups of samples. The box plot analysis provides a visual representation of raw (a) and normalized (b) expression values. RNA-sequencing sample distance analysis (c): RNA-sequencing count tables were statistically analyzed with DESeq2 (Love, Huber, and Anders 2014), and the Euclidean distances were calculated between each sample; samples were clustered using hierarchical clustering analysis, and the dendrograms represent the clustering results. The heatmap illustrates the pairwise distances between the indicated samples, with the colors indicating the distances as shown in the key in the right; i.e., the more blue the square, the more similar the samples). (d) Principal component analysis: PC1 and PC2 variance of expression between RNA-seq samples is shown. The plot shows the projection of the samples onto the two-dimensional space spanned by the first and second principal components of the covariance matrix. The expression levels used as input are normalized log CPM values.

We found 1035 muscular differentially expressed genes (DEGs) between our patient and controls (fig. 7a). The hierarchical clustering analysis of gene expression data confirmed a highly similar expression profile for which controls clustered together apart from the *CCDC78* mutated patient (fig. 7b).

**Fig. 7.**
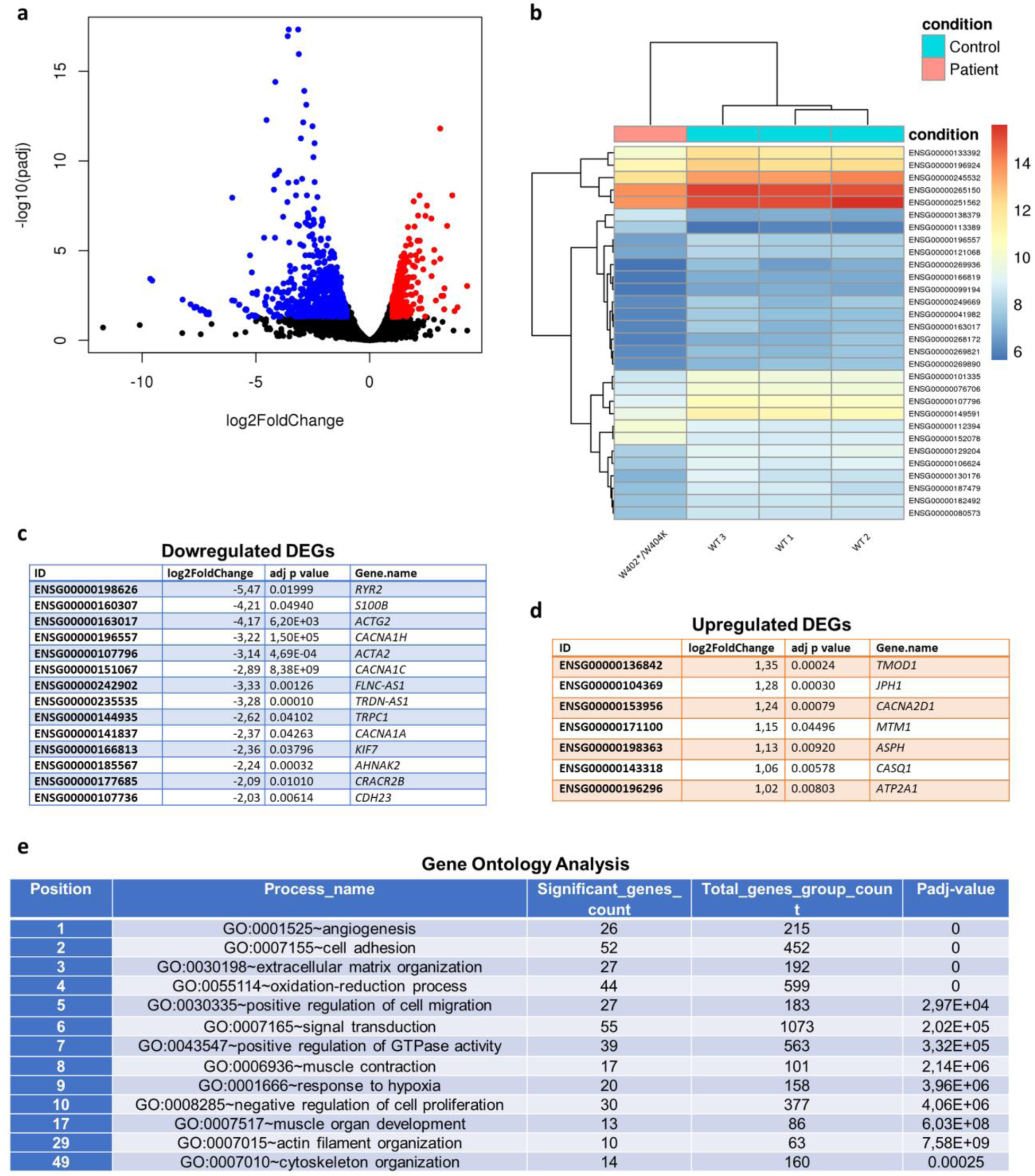
Volcano plots showing gene expression differences between the CCDC78 mutated patient and controls. **(a).** The plot shows the global transcriptional change across the groups compared. All the genes are plotted, and each data point represents a gene. The log2 fold change of each gene is represented on the x-axis and the log10 of its adjusted p-value is on the y-axis. Genes with an adjusted p-value less than 0.05 and a log2 fold change greater than 1 are indicated by red dots and represent upregulated genes. Genes with an adjusted p-value less than 0.05 and a log2 fold change less than -1 are indicated by blue dots and represent downregulated genes. **Hierarchical clustering analysis of the CCDC78 mutated patients and controls with heatmap density color representation of differentially expressed genes (DEGs) (b).** This analysis is performed to visualize the expression profile of the top 30 genes sorted by their adjusted p-values. This analysis is useful to identify co-regulated genes across the conditions. In **c** and **d** are reported a series of downregulated and upregulated genes associated with muscular function and SR. **Gene ontology (GO) analysis (e): t**op GO terms of genes associated with differentially expressed transcripts identified in muscles from the *CCDC78* mutated patient as compared to controls. P-values were adjusted by false discovery rate (FDR) multiple testing correction.

To assess the biological functions of the identified *CCDC78* patient associated DEGs, we performed a functional gene enrichment analysis. In the discovery set, we observed a statistically significant association with different biological processes. Among them, genes associated to “*extracellular matrix organization”* have been previously proved differentially expressed in other types of genetic myopathy^11^ and seems to be common to many dystrophic processes. The top 10 processes sorted by their adjusted p-values were: *angiogenesis*, *cell adhesion*, *extracellular matrix organization*, *oxidation-reduction process*, *positive regulation of cell migration*, *signal transduction*, *positive regulation of GTPase activity*, *muscle contraction*, *response to hypoxia* and *negative regulation of cell proliferation* (fig. 7e). Interestingly, among all the 411 statistically significant processes sorted by their adjusted p-values and most relevant to a muscle disease we found the following: *muscle contraction* (8th position), *muscle organ development* (17th), actin filament organization (29th), *cytoskeleton organization* (49th), *calcium ion transport into cytosol* (60th), *response to muscle activity* (70th), *actin cytoskeleton organization* (77th), *detection of calcium ion* (107th), *actomyosin structure organization* (132th), *calcium ion transport* (176th), *clustering of voltage-gated sodium channels* (179th), *endoplasmic reticulum organization* (266th), *actin cytoskeleton reorganization* (284th), *actin filament bundle assembly* (289th), *membrane depolarization during action potential* (290th), *calcium ion transmembrane transport via high voltage-gated calcium channel* (311th), *negative regulation of skeletal muscle cell differentiation* (315th), *calcium ion import* (361th).

Among the 1035 muscular DEGs, we found upregulation with a log2 fold change ranging from 1.02 to 1.35 of a series of genes involved in SR and excitation–contraction coupling (ECC): *TMOD1, JPH1, CACNA2D1, MTM1, ASPH, CASQ1* and *ATP2A1* (fig. 7d). Among the other top upregulated DEGs we found the following: *PKP2, DBNDD1, ARHGAP36, MSTN, MYLK2, SGCD, TTL, VEZT, WHAMM, SEMA4D, NIN* and *PLXNA1.* Among the top downregulated DEGs we found the following: *S100B, ACTG2 CACNA1H, ACTA2, CACNA1C, FLNC-AS1, TRDN-AS1, TRPC1, CACNA1A, KIF7, AHNAK2, CRACR2B, CDH23* (fig. 7c).

### 4. CCDC78 interacts with two pivotal SR proteins: SERCA and CASQ1

Our Co-Ip assays clearly showed the presence of several bands (fig. 8a), each corresponding to a potential different CCDC78 interacting protein. A band located between 37 and 50 kDa corresponding to CCDC78 was identified in the CCDC78 pulldown lane or input lane but not in the IgG control lane on the blue Coomassie gel and WB: this finding demonstrates that CCDC78 can be efficiently and specifically immunoprecipitated from the muscle tissue extract. In the blue Coomassie gel we identified a band corresponding to ∼110 kDa (band 2) and a ∼50 kDa band (band 4); they were manually excised from the gel for the nLC-nESI-HRMS/MS analysis, as described in material and methods section. From the band 2, applying the filtration criteria described in the experimental procedure and considering a significance threshold p<0.05, we were able to identify 60 database entries. By filtering on total score, emPAI and mass ≥100 kDa and ≤120 kDa, we were able to identify sarco-endoplasmic reticulum calcium ATPase1 (SERCA1), having a sequence coverage of 28% (fig. 8b-c, Table S1). Band 4 was analyzed according to the same procedure for band 2; we thus identified 111 database entries, and, among these, we selected calsequestrin1 (CASQ1), having a sequence coverage of 11% (fig. 8b-d). We then confirmed these findings by WB analysis. Notably, the presence of the CCDC78 mutation was able to abolish the interactions with SERCA1 and apparently to decrease the interaction with CASQ1 (fig. 8e). Our results were corroborated by our transcriptome data that showed an upregulation of both *ATP2A1* and *CASQ1* in our *CCDC78* mutated patient compared to controls (fig. 7d).

**Figure 8.**
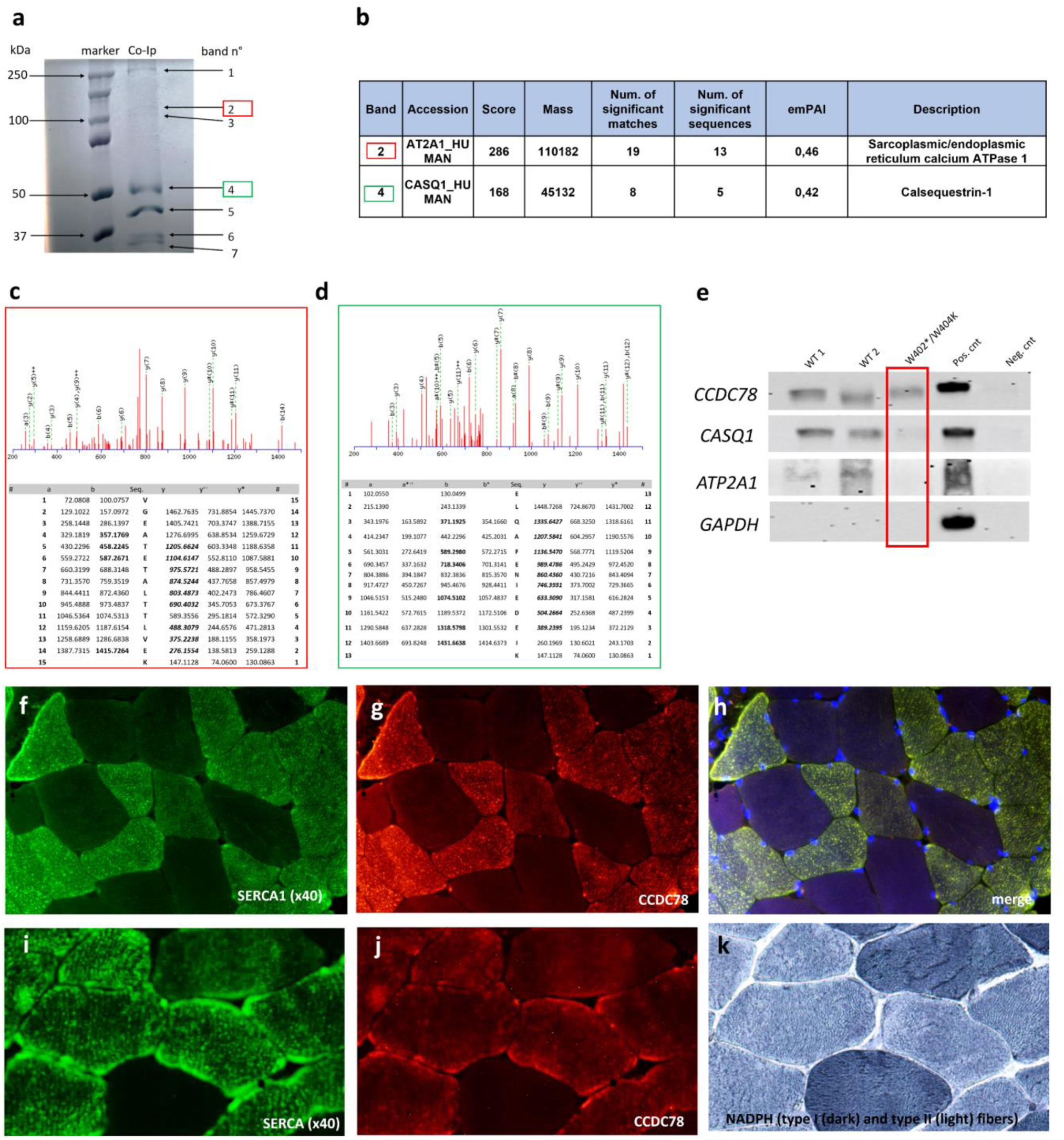
Co-immunoprecipitation (Co-Ip) of CCDC78-interacting proteins. **(a)**: muscle lysates were subjected to Co-Ip using anti-CCDC78 antibody and analyzed by SDS-PAGE followed by Coomassie blue staining. Two bands with a molecular weight of around 50 and 110 kDa were detected (band 2, in red, and 4, in green). Lane 1, protein molecular weight ladder; lane 2, muscle lysate of healthy control. **Identification of CCDC78 interacting proteins by nLC-nESI-HRMS/MS (b)**. **Mass spectrometry (MS) analysis of bands 2 (c)**: fragmentation spectra of 781.420532,2 m/z identifying AT2A1_HUMAN. The corresponding putative aminoacid sequences is taken from MASCOT search. The VGEATETALTTLVEK producing an Ions Score of 52 (expect: 5.9e-005). The m/z values of detected positive ion fragments are in red. The ‘b’ and ‘y’ ions are singly charged fragments (molecule + 1 H +) produced by fragmentation from the N- and C-terminus respectively; ‘b ++ ’ and ‘y ++ ’ ions are the corresponding doubly charged fragments (molecule + 2 H +). The y* ions are y ions with loss of water. **MS analysis of bands 4 (d)**: fragmentation spectra of 789.388794 m/z identifying CASQ1_HUMAN and the corresponding putative aminoacid sequences taken from MASCOT search. The ELQAFENIEDEIK producing an Ions Score of 47 (expect: 3.9e-005). **Western blot analysis (e)**. Proteins immunoprecipitated using CCDC78 antibody were immunoblotted on membranes using anti-ATP2A1 and anti-CASQ1 antibodies. ATP2A1 and CASQ1 were detected in wild-type pulldowns (WT1, WT2) but not in CCDC78 mutated patient pulldown (PT). ATP2A1 and CASQ1 were also detected in the total cell lysate (+Cnt) but not in the IgG control (-Cnt.). Membranes were immunoblotted with anti-GAPDH (negative Co-IP control) and anti-CCDC78 (positive Co-IP control). **Colocalization between SERCA1 and CCDC78 in control muscle tissue (f-h)**. In the muscle seriated sections, we found a colocalization of CCDC78, SERCA1 and NADPH diaphorase **(i-k)**.

Immunofluorescent study in muscle tissue was also highly consistent with the found proteins interactions; we indeed observed a stringent colocalization between CCDC78 and SERCA1 (fig. 8g-h) in control muscle tissue. Notably, in the histological serial sections we found a colocalization of CCDC78, SERCA1 and NADPH diaphorase, thus confirming the predominantly localization of CCDC78 in type II fibers (fig. 8i-k). In the CCDC78 mutated muscle some SERCA1 (fig. 9) and CASQ1 aggregates were evident. Notably, in our *RYR1* mutated patients showing central cores, we observed a costaining of SERCA1, CCDC78 (fig. 9g-i) and CASQ1 aggregates in correspondence of cores.

**Figure 9.**
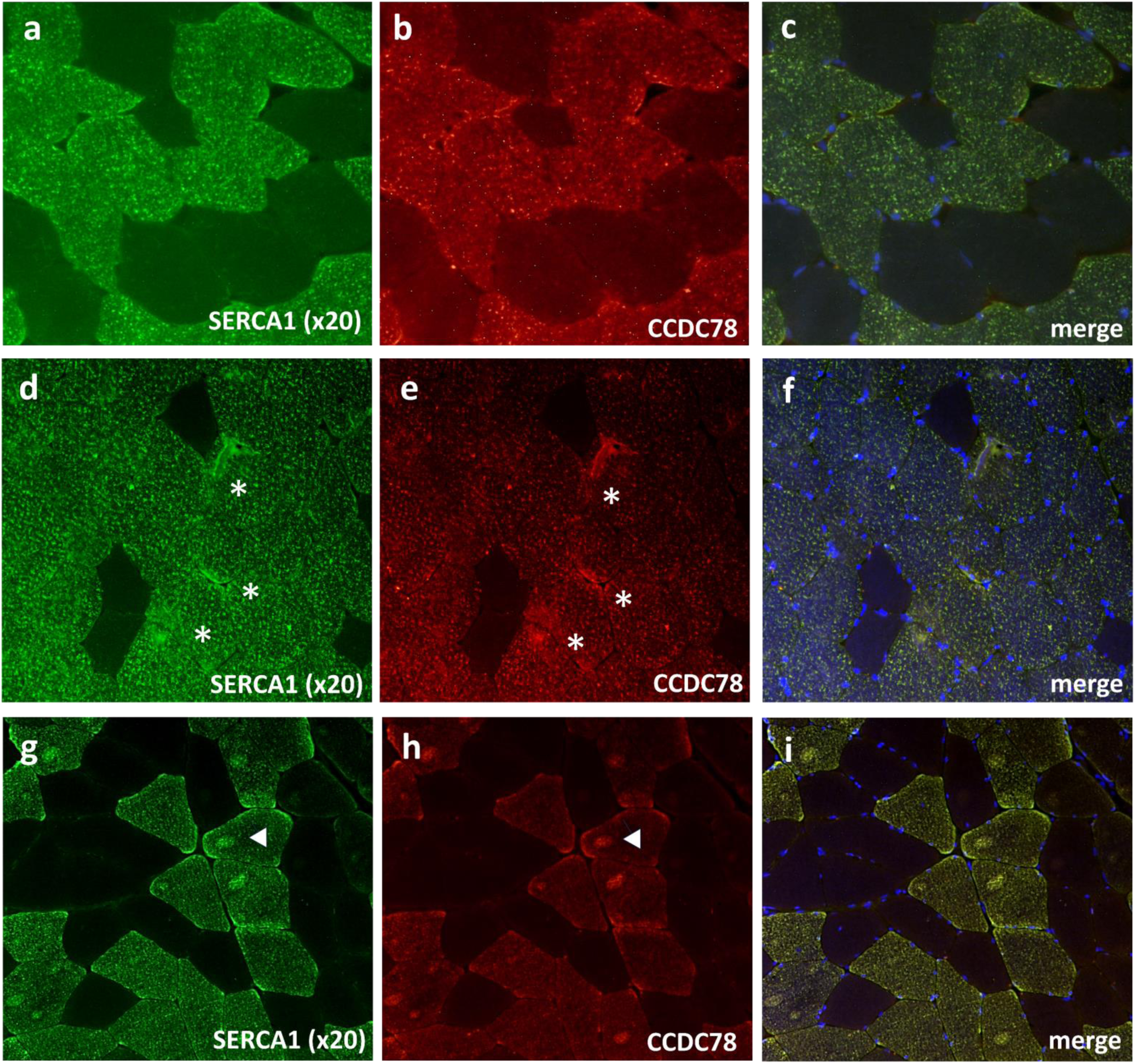
SERCA1 and CCDC78 aggregates (asterisks) in *CCDC78* mutated muscle (d-f) compared to control (a-c). RYR1 mutated muscle (g-i): costaining of SERCA1 and CCDC78 with cores (arrow heads).

### 5. CCDC78 interacts with other proteins outside the SR: MYH1, ACTN2, ACTA1

In the blue Coomassie gel we also identified three other bands corresponding to ∼250kDa (band 1), ∼100kDa (band 3) and 37-50kDa (band 5), respectively. These bands underwent the same procedures as above for nLC-nESI-HRMS/MS analysis. From band 1 we were able to identify 168 database entries, and, after the filtering analysis, we selected MYH1, having a sequence coverage of 58% (fig. 10a-c, Table S1). As for band 3 and 5, we identified respectively 73 and 56 database entries and, among these, we selected ACTN2, having a sequence coverage of 39%, and ACTA1, having sequence coverage of 63% (fig. 10a,b,d). We then confirmed by WB the interaction of CCDC78 with MYH1, ACTN2 and ACTA1 in the analyzed muscle samples. Even in this case the *CCDC78* mutation seems to decrease the interaction between CCDC78 and ACTA1 (fig.10,f-g).

**Fig. 10.**
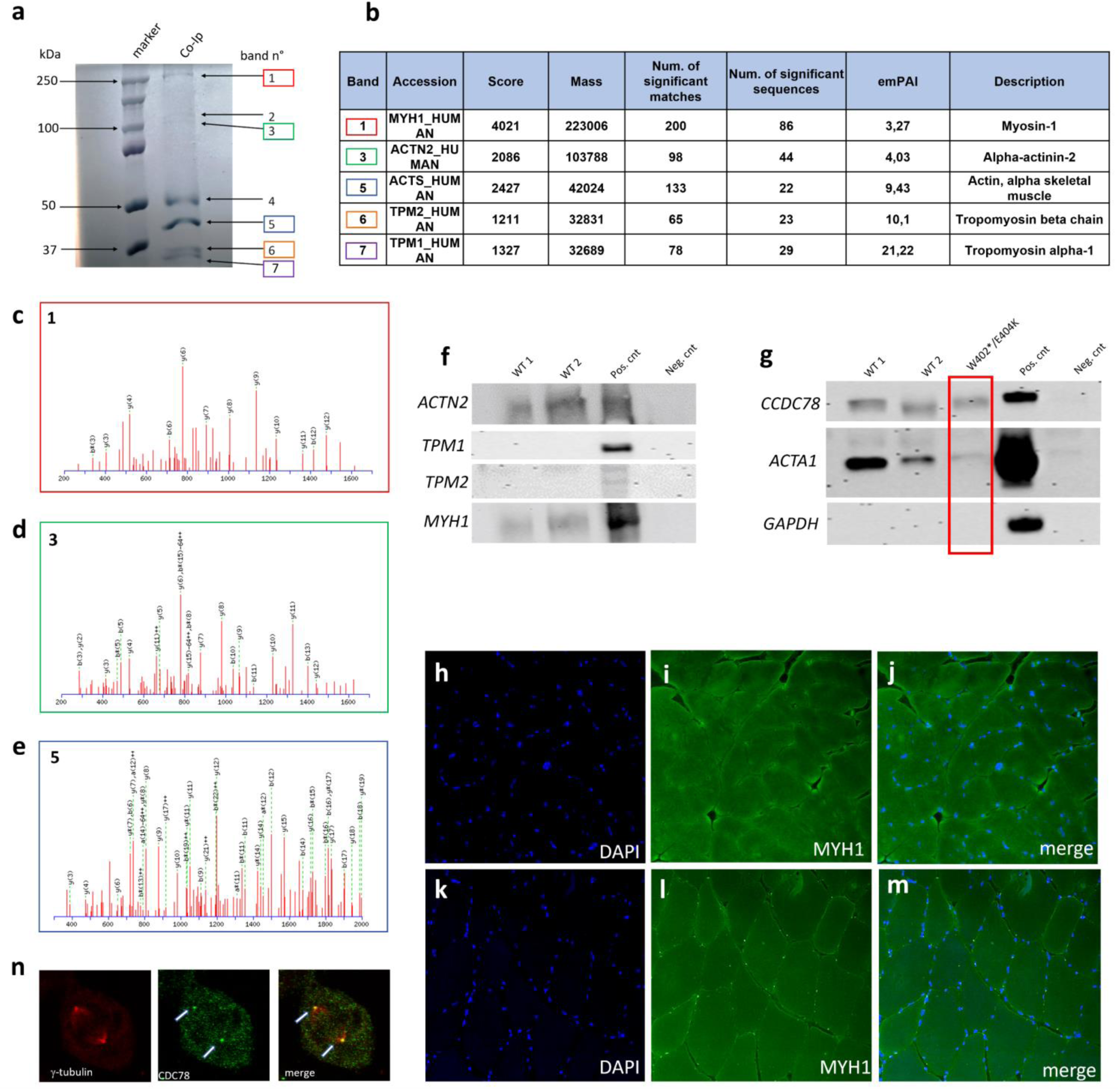
Co-Ip of CCDC78-interacting proteins. Five bands were futher detected: ∼250 kDa (band 1, in red), ∼100 kDa (band 3, in green), ∼45 kDa (band 5, in blue), two bands in the 25-37 kDa range (band 6, in orange, band 7 in purple). Lane 1, protein molecular weight ladder; lane 2, muscle lysate of healthy patient. **Identification of CCDC78 interacting proteins by nLC-nESI-HRMS/MS (b)**. **Mass spectrometry (MS) analysis of band 1 (c):** fragmentation spectra of 859.917908 m/z identifying MYH1_HUMAN. The LQNEVEDLMIDVER sequence produced an Ions Score of 81 (expect: 6.7e-008). **Mass spectrometric analysis of bands 3 (d)**: fragmentation spectra of 908.426208 m/z identifying ACTN2_HUMAN. The ISSSNPYSTVTMDELR sequence produced an Ions Score of 82 (expect: 2.1e-008). **Mass spectrometric analysis of bands 5 (e):** fragmentation spectra of 1276.581421 m/z identifying ACTS_HUMAN. The LCYVALDFENEMATAASSSSLEK sequence produced an Ions Score of 77 (expect: 5.6e-008). **Western blot analysis (f-g).** Proteins immunoprecipitated using CCDC78 antibody were immunoblotted using anti-ACTN2, anti-MYH1, anti-ACTA1, anti-TPM1 and anti-TPM2 antibodies. ACTN2, MYH1 and ACTA1 were detected in wild-type pulldowns (WT1, WT2). ACTN2, MYH1 and ACTA1 were also detected in the total cell lysate (CTR+) but not in the IgG control (CTR-). Membranes immunoblotted with anti-TPM1 and anti-TPM2 detected protein only in the total cell lysate (CTR+) but not in the WT muscles and IgG control (CTR-). Membranes were immunoblotted with anti-GAPDH (negative Co-IP control) and anti-CCDC78 (positive Co-IP control). **MYH1 staining in control (h-j) and CCDC78 mutated (k-m) muscles (20x):** perinuclear small MYH1 aggregates are present in mutated muscle. HeLa cells showing a costaining of CCDC78 and gamma-tubulin **(n).**

The nLC-nESI-HRMS/MS analysis of band 6 and 7 (fig.10a) identified 70 and 68 entries, respectively. By filtering on total score, emPAI and mass ≥25kDa and ≤37kDa, we selected TPM1 and TPM2 as possible interactors (fig. 10b), however WB did not confirm such further possible interactions (fig. 10f).

Our immunofluorescent studies in muscle tissue showed perinuclear small MYH1 aggregates (fig. 10h-m) and a small number of ACTA1 aggregates in our *CCDC78* mutated patient compared to controls. Other than the above selected proteins, we analyzed in HeLa cells the possible colocalization of CCDC78 and gamma-tubulin, since the previous association of CCDC78 with centriolar structures: we found a localization of both the proteins at the spindle pole (fig. 10n), the region of the cell where the centrosome is located, thus supporting a role of CCDC78 in centrioles.

### 6. CCDC78 does not interact directly with RyR1

Our *CCDC78* mutated patient showed muscular RyR1 aggregates colocalizing with CCDC78, thus confirming the previous data of Majcenko et al^8^; interestingly, in CCDC78 mutated muscle, RyR1 aggregates co-stained with DAPI both in peripheral and central regions of the fibers thus suggesting a localization of RyR1 aggregates in a perinuclear reticulum region (fig. 11e-f). We also observed Trisk95 aggregates colocalizing with RyR1 (fig. 11e-f). The morphometric analysis of muscle fiber cross-sectional area revealed in our *CCDC78* mutated patient a significant increase in % of positive nuclei for RyR1 (16±3% vs 1±0.5% using monoclonal anti-RyR1 antibodies; 15.5±2% vs 2±1% using polyclonal antibody) and Trisk95 (33±3% vs 3±1%) compared to controls (fig. 11d). However, our CCDC78 Co-Ip assay did not reveal any band corresponding to the molecular weight expected for RyR1. Moreover, our confirmation WB study using anti-RyR1 antibodies after CCDC78-Ip did not reveal any interaction between such proteins (fig. 11c).

**Fig. 11.**
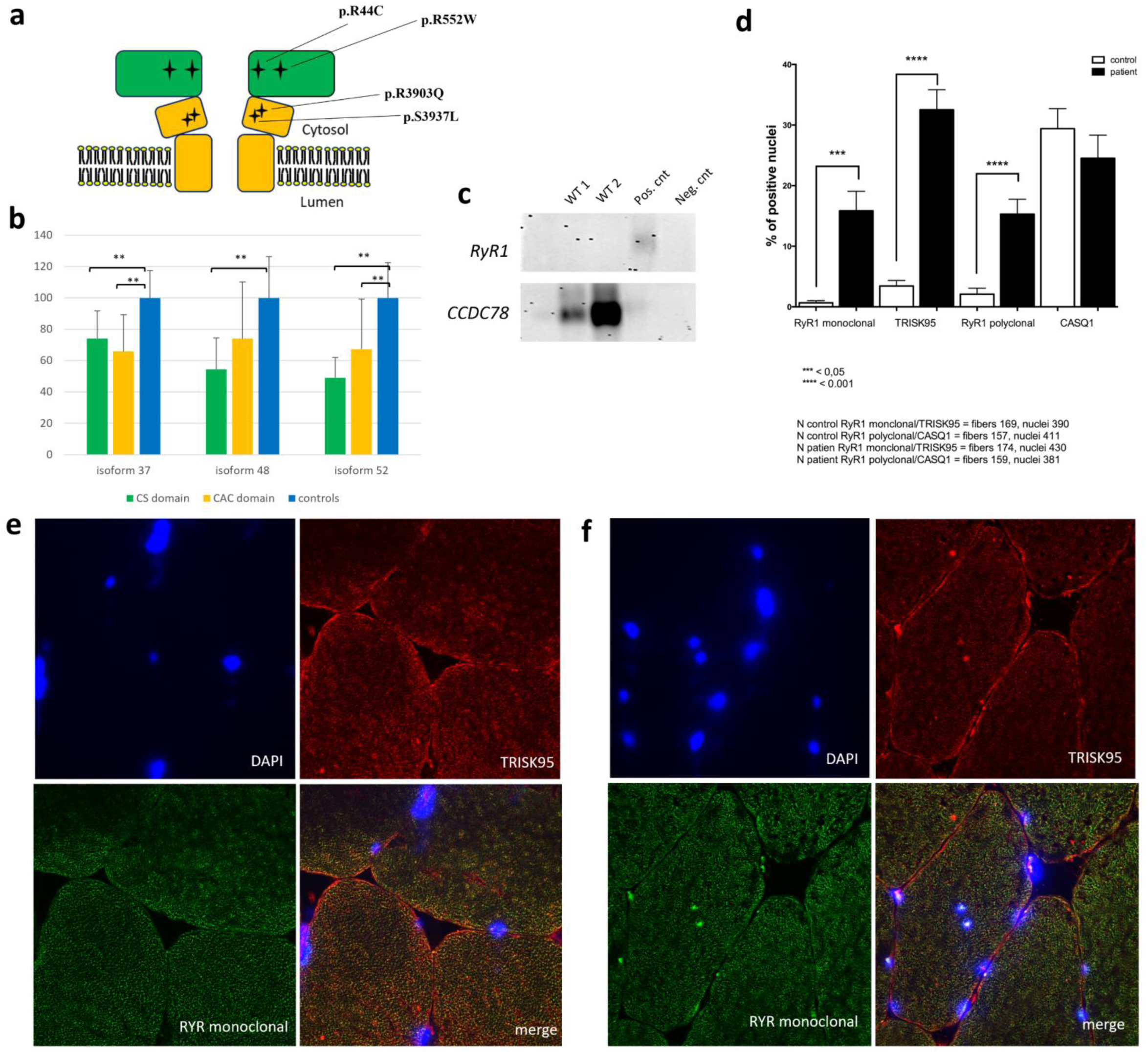
Schematic representation of cytosolic shell (CS, in green) and channel and activation core (CAC, in yellow) of RyR1. **(a):** black star signs represent the analyzed *RYR1* mutations. CCDC78 expression analysis by WB in *RYR1* mutated patients (n=4) compared to controls (n=3) (b): significantly reduction (p.<0.01) of 37 kDa, 48 kDa and 52 kDa isoforms compared to controls. The two *RYR1* mutated patients harbouring a mutation in the CS showed a significantly reduction in 37, 48 and 52 kDa isoforms compared to controls (p<0.01); the patients carrying a *RYR1* mutation in CAC showed a significantly reduction only for 37 and 52 kDa isoforms. **Western blot analysis (c)**: Proteins immunoprecipitated using CCDC78 antibody were immunoblotted using anti-RyR1 and anti-CCDC78 antibodies. CCDC78 was detected in wild-type pulldowns (WT1, WT2), in the total cell lysate (positive control) but not in the IgG control (negative control). Membranes immunoblotted with anti-RyR1 detected the protein only in the total cell lysate. **Morphometric analysis of muscle fiber cross-sectional area (d)**: in *CCDC78* mutated muscle we observed a significant increase of %positive RyR1 and Trisk95 nuclei compared to controls. **RyR1, DAPI and Trisk95 staining in the CCDC78 mutated muscle (f) and control (e)**: in mutated muscle RyR1 aggregates co-stained with DAPI both in peripheral and central regions of the fibers; we also observed Trisk95 aggregates that colocalized with RyR1.

Muscular CCDC78 expression analysis by WB in *RYR1* mutated patients (n=4) compared to healthy controls (n=3) showed a significant reduction of the 37-kDa, 48-kDa and 52-kDa isoforms compared to controls (respectively 70.45% (SD±20.15%, p<0.01), 62.93% (SD±29.03%, p<0.01), 57.05% (SD ± 24.82%, p<0.05)). Interestingly the two *RYR1* mutated patients harbouring a mutation located in the cytosolic shell (CS) of the protein (c.130C>T (p.Arg44Cys) and c.1654C>T (p.Arg552Trp)) showed a significant reduction of the 37, 48 and 52-kDa isoforms (respectively 73.95% (SD±17.93%, p<0.01), 54.34% (SD±20.2%, p<0.01)) and 49.05% (SD±12.99%, p<0.01) while the two *RYR1* mutated patients carrying a mutation in channel and activation core (CAC) (c.11810C>T (p.Ser3937Leu) and c.11708G>A (p.Arg3903Gln)) showed a significant reduction only in 37- and 52-kDa isoforms (respectively 65.94% (SD±23.32%, p<0.01), 67.33% (SD±31.98%, p<0.01)) (fig.11a-b). Muscular RyR1 expression analysis by WB in the *CCDC78* mutated patient did not reveal significant differences compared to controls.

## Discussion

Our data, obtained by deeply analyzing a family harbouring the first *CCDC78* nonsense mutation, allow shading light on the CCDC78 function by defining its major interacting proteins and allow us to expand the muscular phenotype previously associated to the gene. We thus definitely associate this candidate gene to the autosomal dominant centronuclear myopathy-4 (CNM4; OMIM#614807). Our Co-Ip assays, followed by mass spectrometry study and a validation WB phase, showed an interaction between CCDC78 and two pivotal SR proteins: SERCA1 and CASQ1. The SR represents a specialized form of the endoplasmic reticulum of muscle cells, involved in Ca2+ regulation. The Ca2+ removal from the cytosol following muscle contraction is mainly due to SERCA1 that actively pumps Ca2+ from the cytoplasm to the SR lumen^12^. The triad, the structure composed by two terminal cisternae (TC) and one transverse tubules (TT), supports the ECC in skeletal muscle fibers by allowing the interaction between the dihydropyridine receptor (DHPR) and RyR1^13^. At triads, several proteins participate in the ECC process, including CASQ1, a luminal Ca2+ binding protein that interacts with RyR1, triadin, and junctin, forming a quaternary Ca2+ release complex^12^. CASQ1 thus represents the main Ca2+ buffer in the TC^14^ and it has been proposed that its depolymerization, during sustained SR Ca2+ depletion, may represent the intracellular switch that induces RyR1 closing^15^. Recently, it has been demonstrated that CASQ1 acts as a regulator of Store Operated Calcium Entry (SOCE) pathway by binding to STIM1. In particular, CASQ1 overexpression was found to inhibit SOCE by reducing STIM1 clustering and STIM1/OraiI interaction.

The interaction of CCDC78 with SERCA1 and CASQ1 that we found highly supports the role of CCDC78 in SR and therefore in ECC. Interestingly, we showed that the presence of the *CCDC78* mutation determined a loss or reduction of interaction between these proteins thus suggesting a possible interaction site between such proteins. According to the role of CASQ1, SERCA1 and other related SR proteins, the transcriptome analysis in *CCDC78* mutated muscle revealed the presence of peculiar pattern of differentially expressed genes. Indeed, among the most upregulated genes, we found a series of genes involved in SR and ECC: *TMOD1, JPH1, CACNA2D1, MTM1, ASPH, CASQ1* and *ATP2A1*. Upregulation of *CASQ1* and *ATP2A1*, encoding SERCA1, is highly suggestive of a compensatory effect due to the *CCDC78* mutation and further supports the intimate role of such *CCDC78* interactors.

Tmod1, encoded by *TMOD1*, may be associated with a subdomain of the RyR1-containing SR compartment flanking the Z line^16^. Junctophilin 1, encoded by *JPH1* and located at the triad, mediates the apposition between ER/SR and plasma membrane in striated muscles; it can also bind and regulate various proteins at triads, acting as a scaffold for assembly of the Ca2+ release complex^17, 18^. *CACNA2D1* encodes the α2-δ subunit of the DHPR, a voltage-gated calcium channel and voltage sensor for Ca2+ release in skeletal muscle^19^. *MTM1* encodes for myotubularin, a lipid phosphatase localized at triads whose balanced expression levels are required for the proper assembly of SR and TT into triads^12^; MTM1 may also be involved in regulating membrane curvature by acting on PtdIns3P-rich membrane subdomains^20^. The *ASPH* gene encodes 3 proteins, aspartate beta-hydroxylase, junctin, and junctate: junctin is structurally similar to triadin and, like triadin, can bind CASQ1 and RyR1^12^; junctate is a Ca2+-sensing structural component of the Endoplasmic reticulum (ER)-plasma membrane (PM) junctions where Orai1 and STIM1 cluster and interact to mediate SOCE^21^.

Even among the top downregulated DEGs we found some genes also involved in SR: *CRACR2A* and *TRDN-AS1*. CRACR2A regulates SOCE by associating in a ternary complex with ORAI1 and STIM1, both critical components of calcium release-activated calcium channels involved in SOCE into cells^22^. Moreover, CRACR2A has a role in enhancement of Orai1-mediated SOCE. Thus, CRACR2A shows an opposite function compared to CASQ1; the *CRACR2A* downregulation corresponds indeed to the CASQ1 upregulation observed in our *CCDC78* mutated muscle. *TRDN-AS1* (Triadin Antisense RNA 1) is an RNA gene affiliated with the lncRNA class^23^. *TRDN-AS1* overlaps with skeletal muscle *TRDN* and is expressed in an opposite orientation to regulate the balance between the cardiac and skeletal muscle isoforms of *TRDN*. In absence of *TRDN-AS1* transcription, *TRDN* is indeed fully transcribed, giving rise to the skeletal muscle isoform of *TRDN* (Trisk95 and Trisk51)^24^. Triadin acts as a functional regulator of EEC by interacting with RyRs, junction, CASQ1 and the histidine-rich Ca2+-binding protein^12^. Triadin may also play a structural role in supporting triad architecture by interacting with *CKAP4*. Thus, the downregulation of *TRDN-AS1* observed in our *CCDC78* mutated muscle may drive a consequent *TRDN* upregulation.

This finding is in line with our immunofluorescence analysis, showing a significant increase of Trisk95 positive nuclei in mutated muscle fibers compared to control ones.

The observed reduction of CCDC78 muscular expression by WB in our RyR1 mutated patients, especially when mutations are located in the CS of RyR1, may further support the interaction between CCDC78 and CASQ1, since CS represents the interacting domain of RyR1 with triadin, junctin, and CASQ1^25, 26^. The interaction of CCDC78 with CASQ1 and SERCA1 allows concluding that CCDC78 localizes into SR and supports a possible role of the protein in connecting these two pivotal SR proteins, thus stabilizing the SR and contributing to ECC processes. Notably, the observed SR dilatations in *CCDC78* mutated muscle corroborate the role of CCDC78 in this cell compartment. Since the protein has been demonstrated to interact with multiple sarcoplasmic proteins, we suggest renaming it *sarcoplasmin* and *SRPM* the related gene.

The CCDC78 involvement in the triad was previously hypothesized by Majczenko et al^8^ that showed by immunostaining a reticular pattern partially overlapping the triad. Moreover, in mutated patients, they showed that CCDC78 accumulated in large sarcoplasmic aggregates, which co-stained for desmin, actin, RyR1 and DHPR1α. Our immunostaining study confirms the previous findings of CCDC78 co-localization with RyR1 and desmin in muscular aggregates. Surprisingly, we did not detect an interaction between RyR1 and CCDC78; this was in line with our transcriptome findings, showing no differences in RyR1 expression between *CCDC78* mutated and controls muscles. However, the presence of RyR1 aggregates in *CCDC78* mutated muscle, the significantly reduction of different CCDC78 isoforms and the CCDC78 aggregates colocalizing with cores in RyR1 mutated patients, together with the well-known interaction of RyR1 with CASQ1^12^, likely suggest a possibly indirect interaction between RyR1 and CCDC78. However, more studies are needed to further explore such hypothesis.

Our data, showing the interaction of CCDC78 with CASQ1 and SERCA1, both proteins largely predominant in fast-twitch (type II) fibers^27, 28^, together with the results from NADPH diaphorase, lead to conclude that CCDC78 is mainly localized in fast-twitch (type II) fibers. Interestingly, our mutated patient showed predominance of type II fibers on muscle biopsy, in line with the observed higher CCDC78 expression in mutated muscle tissue compared to controls, probably due to a compensatory effect.

Other than the above-described interactions with some SR proteins, we also found an interaction of CCDC78 with the following extra-SR proteins: alpha-actin (ACTA1), alpha-actinin-2 (ACTN2) and myosin-1 (MYH1). ACTA1, the predominant actin isoform in the sarcomeric thin filaments of skeletal muscle, is essential, along with myosin, for muscle contraction. ACTA1 interacts with the other thin filament proteins including nebulin, tropomyosins and troponins in the Z-disc, and importantly with myosins in the thick filament^29^. Although *RYR1* mutations are the main genetic cause of multiminicore disease and core rod myopathy^30–32^, characterized by central cores and nemaline bodies (i.e. rods, mainly composed of actin and α-actinin^33^), even *ACTA1* mutations have been rarely associated to such conditions^34^. The found interaction between CCDC78, involved in SR, and ACTA1, whose mutations are sometimes associated to a RYR1-like muscular phenotype, may suggest a possible role of CCDC78 in connecting ACTA1 to SR structures. This hypothesis is reinforced also by the interaction of CCDC78 with ACTN2. ACTN2 is indeed an actin-binding protein that crosslinks actin across the entire Z-disk^35, 36^. It is involved in sarcomere formation, anchoring/crosslinking of actin thin filaments, and interaction with titin^36, 37^. Moreover, α-actinins have been proven to interact directly or indirectly with elements of intermediate filaments, SR, costameres, and signaling molecules^38^.

Also MYH1 represents a cytoskeletal muscular protein interacting with actin^39^. Although MYH1 has not been clearly associated to date with *RYR1*-related myopathies, *MYH1* mutations were associated with non-exertional rhabdomyolysis phenotype in horses^40^ and *MYH1* was recently considered a candidate gene for recurrent rhabdomyolysis also in humans^41^. Interestingly, the proteomic study of Eckhardt et al^42^ showed a quantitative reduction of MYH1 in a mouse model with biallelic *RYR1* mutations; accordingly, Wang et al^43^ showed a significant reduction of *MYH1* expression in muscle tissue from *RYR1* mutated patients and suggested MYH1 as potential biomarkers for *RYR1-*myopathies. These data, together with the found interaction with CCDC78, reinforce the hypothesis of an intimate role of MYH1 with other SR proteins.

A muscular clinical phenotype associated to *CCDC78* mutations was previously described in only one family by Majczenko et al.^8^: the phenotype included early-onset distal muscle weakness, myalgia, easy fatigue, mild cognitive impairment and normal CPK levels. Differently from Majczenko et al., our *CCDC78* mutated patient showed adult onset myalgias and painful muscle cramps with bilateral calf hypertrophy and persistent moderate CPK elevation; the patient’s relative, harbouring the same mutation, was asymptomatic at 18-25 years of age and showed only bilateral calf hypertrophy. No cognitive impairment was detected in our patients. Although the presence of the already described nuclear centralizations^8^ in muscle biopsy, our patient did not show any core-like lesions differently from the previous study; moreover, ultrastructural analysis revealed peculiar SR abnormalities previously described only in the ccdc78 zebrafish morphants harbouring a splice-site mutation; indeed, in humans, only distension of transverse (T)-tubules was previously described^8^. These data may suggest a more prominent muscle involvement in the presence of nonsense variants suggesting a likely haploinsufficiency effect.

However, we cannot exclude that such slightly different phenotype could be mediated by the different variant effect on different *CCDC78* isoforms. Indeed, in different control tissues including muscle, we surprisingly found, other than the presence of the two already known *CCDC78* transcripts (NM_001031737.3 and NM_001378030.1), two alternative in-frame transcripts deriving from alternative splicing in the last exons. As previously demonstrated^44^, through alternative pre-mRNAs processing, individual mammalian genes often produce multiple mRNA and protein isoforms that may have related, distinct, or even opposing functions. It is thus possible that CCDC78 products with different length may act different role in muscle and different tissues and, consequently, different mutations could lead to a kaleidoscopic effect on phenotypes. This hypothesis is in line with our findings of a series of CCDC78 interactors inside and outside the SR, that highly indicates a role in different cellular compartments.

We provided for the first-time evidence that a nonsense *CCDC78* variant results in a mutant allele with premature stop codon, which is likely mostly degraded on mRNA level by means of NMD. However, we observed 84% of 48kDa-isoform expression in the mutated patient compared to controls, higher than the expected 50%, possibly indicating some degree of expression compensation by increased wild-type translation. Moreover, our WB study revealed a significant 52kDa isoform reduction in the mutated patient; since the mutation results as a missense variant (c.1210G>A, p.Glu404Lys) in the NM_001378030.1 transcript, such reduction probably reflects a dominant negative effect driven by the missense variant. Missense mutations can indeed affect DNA-transcription factors resulting in altering the expression of the corresponding protein; beside the fact that reduced transcription or unstable mRNA can lead to reduced protein expression, disease-causing missense mutations are thought to cause intracellular retention of their respective mutant proteins^45, 46^.

Among the significant processes from GO analysis we found *“muscle contraction”*, *“calcium ion transport into cytosol”*, *“response to muscle activity”* and “*endoplasmic reticulum organization”,* thus supporting the prominent role of CCDC78 in the ECC. However, we also found a series of significant processes involved in cell division, cytoskeleton organization and therefore in myogenesis: *“positive regulation of cell migration”*, *“negative regulation of cell proliferation”*, *“muscle organ development”*, *“actin filament organization”*, *“cytoskeleton organization”*, *“negative regulation of skeletal muscle cell differentiation”*. Klos Dehring et al^47^ demonstrated in a multi-ciliated cell model that CCDC78 represents a centriole-associated and deuterosome protein that is essential for amplification of centrioles, microtubule-based structures involved in centrosomes, cilia, and flagella formation. Although primary cilia have long been identified in skeletal muscle, their role remains to date not well understood. However, some recent studies indicate a possible role of cilia in muscle progenitors where they regulate cell cycle progression, trigger differentiation and maintain a commitment to myogenesis^48^. Recently, Palla et al^49^ found that the ability of muscle stem cells to regenerate is regulated by the primary cilium, a cellular protrusion comprised of microtubule bundles organized into an axoneme and anchored by a mature centriole or basal body. Our RNA-seq results, indicating a possible role of *CCDC78* in cell division process, and the colocalization of CCDC78 and gamma-tubulin in HeLa cells during cell division, highly suggest a possible role of the protein in muscle cell division.

Moreover, among the top downregulated DEGs we found some genes involved in cytoskeleton organization: *AHNAK2, CDH23, KIF7.* AHNAK2 is a costameric protein showing co-localization with α-actinin in a region at the end of the Z-disks^50^; CHD23 is a member of the cadherin superfamily, that seems to be involved in stereocilia organization and hair bundle formation^51^; *KIF7* encodes a cilia-associated protein belonging to the kinesin family that plays a role in the hedgehog signaling pathway and in the regulation of microtubule acetylation and stabilization^52^.

## Conclusions

In conclusion, our findings shed light for the first time on interactors and possible role of CCDC78 in skeletal muscle, thus allowing us to definitely locate the protein in SR. Moreover, our data expand the phenotype previously associated with *CCDC78* mutations, indicating new possible histopathologic hallmarks of disease in humans. Future studies including an increasing number of patients harbouring different *CCDC78* mutation are needed to better define the exact associated phenotype and the exact role of CCDC78 in skeletal muscle, both in SR and in extra-SR subcellular structures.

## Materials and methods

### Standard protocol approvals and patient consents

Written informed consent was obtained from all patients involved in the study. All experiments described were approved by the local ethical committee (Regional Ethics Committee for Clinical Trials of the Tuscany Region, n. 17397, approval date 20^th^ July 2020). All the procedures are in accordance with the Helsinki declaration of 1975.

### Morphological analysis of muscle biopsies and HeLa cells

Routine morphology and immunofluorescence analysis of muscle proteins were performed on right rectus femoris muscle biopsy samples according to standard protocols^53^. Ten μm-thick sections were stained with standardized histological and histochemical methods including hematoxylin and eosin, reduced NADH, SDH, and COX. Details for ultrastructural studies are reported in supplementary materials (see supplementary information).

For immunofluorescence experiments, 8 μm-thick sections were incubated with the following primary antibodies: anti-TRISK95 1:500 (Kindly provided by Dr. Isabelle Marty, INSERM, Grenoble, France), Anti-RyR 34C 1:500 (ThermoFisher Scientific), anti-CASQ1 rabbit antibody (ThermoFisher Scientific, Waltham, MA), anti-CASQ1 mouse antibody (clone MA3-913; ThermoFisher Scientific), rabbit anti-CCDC78 (AV53233, Sigma-Aldrich), rabbit Anti-MYH1 (clone 1E15; Sigma-Aldrich), mouse Anti-Actin (alpha-Sarcomeric, clone 5C5, Sigma-Aldrich), and anti-RyR1 rabbit antibody, anti-SERCA1, anti-desmin ^54^. Cy2- or Cy3-conjugated anti mouse or anti rabbit secondary antibodies (Jackson ImmunoResearch Laboratories, Westgrove, PA) were used for immunofluorescence detection.

For immunofluorescence experiments involving HeLa cells we used the protocols already described by Genovese et al^55^ and Vorobjev et al^56^.

Morphometric analysis of muscle fiber cross-sectional area was estimated using Image J^57^. For measurement of fibre cross-sectional area, contrast of micrographs was enhanced by 0.5%. The freehand tool was then used to mark individual fibres.

### DNA analysis

Total genomic DNA was extracted from peripheral blood samples with MasterPure Complete DNA Purification Kit (*Epicentre* MasterPure DNA Purification Kit (cat#: *MCD85201*), according to the manufacturer’s instructions. DNA was analyzed by whole exome sequencing (WES) technique by the NovaSeq6000 (Illumina) on DNA from peripheral blood samples (see supplementary information).

### Cells culture

Peripheral blood lymphocytes (PBLs) were obtained from the CCDC78 mutated patient and one control; mononuclear cells were separated by centrifugation on a Lymphoprep gradient^58^. The cells were washed twice in PBS, resuspended in RPMI 1640 medium with 10% foetal bovine serum (FBS), 1% l-glutamine, 1% penicillin/streptomycin and 1% sodium pyruvate and maintained at 37°C with 5% CO_2_. To assess nonsense-mediated decay (NMD) PBLs were treated with cycloheximide (100 μg per milliliter), an inhibitor of nonsense-mediated decay, for 4 hours. Both PBLs from the patient and the control were collected after 4 h of culture for analysis.

Methods for primary fibroblast cultures and Hela cells^55^ are described in detail in supplementary information.

### RNA extraction

Total RNA, isolated from peripheral blood lymphocytes (PBLs), fibroblasts, and muscle tissue from *CCDC78* mutated patient and controls was processed using RNeasy Mini Kit (Qiagen, Hilden, Germany), following the manufacturer’s instruction. The concentration and purity of total RNA samples were quantified using the Qubit^TM^ RNA IQ assay kit (Thermo Scientific, Massachusetts, USA).

### Transcripts analysis

For each sample, 1 µg of total RNA was reverse transcribed using ImProm-II ™ Reverse Transcriptase (Promega, Wisconsin, USA) and oligodT primers in a 20µl volume. PCR amplifications were performed with primers surrounding the mutated region (supplementary table 2). The products of individual PCR reactions were separated electrophoretically on agarose gels. After the purification of cDNA products with QIAquick PCR Purification kits (Qiagen, Hilden, Germany), sequencing was performed using the automated sequencer ABI 3500 (Applied Biosystems, Massachusetts, USA). The results were analyzed using Chromas software version 2.33 and compared with reference sequence NG_032932.1.

### Transcriptome profiling

Transcriptome profiling by RNA-seq from three control muscle samples and *CCDC78* mutated patient was carried out using the Illumina NovaSeq (San Diego, USA), PE 2x150 via polyA selection. Sequence reads were trimmed to remove possible adapter sequences and nucleotides with poor quality using Trimmomatic v.0.36. The trimmed reads were mapped to the Homo sapiens GRCh37 reference genome available on ENSEMBL using the STAR aligner v.2.5.2b, a splice aligner that detects splice junctions and incorporates them to help align the entire read sequences. Unique gene hit counts were calculated by using featureCounts from the Subread package v.1.5.2. The hit counts were summarized and reported using the gene_id feature in the annotation file. Only unique reads that fell within exon regions were counted. After extraction of gene hit counts, the gene hit counts table was used for downstream differential expression analysis. Reads per exon were grouped, from which FPKM (Fragments Per Kilobase of exon model per Million mapped reads) values will be calculated^59^. Only genes with FPKM >1 were included in further analyses.

Using DESeq2, a well-established and accepted analysis tool^60, 61^, a comparison of gene expression between the defined groups of samples was performed. The Wald test was used to generate p-values and log2 fold changes. Genes with an adjusted p-value < 0.05 and absolute log2 fold change > 1 were called as differentially expressed genes for each comparison. Data quality and sample identity assessment was performed. In particular, the original values were normalized to adjust for various factors such as variations in sequencing amount; these normalized values were used to accurately determine differentially expressed genes.

The significance of the set of genes commonly differentially expressed or spliced was evaluated using a non-parametric test. Gene Ontology (GO) enrichment analysis and pathway enrichment analysis were performed on the statistically significant set of genes by implementing the software GeneSCF v.1.1-p2. The goa_human GO list was used to cluster the set of genes based on their biological processes and determine their statistical significance. A list of genes clustered based on their gene ontologies was generated.

### RT-qPCR

RT-qPCR was performed using QuantiNova SYBR Green RT Mix (Qiagen, Hilden, Germany) according to the manufacturing instructions, with the reaction mixture (total volume 20 µl) containing 20 ng of RNA. All reactions were performed in triplicates on a CFX96 Real Time System (Bio[Rad, California, USA). Light cycler protocol included 10 min of initial reverse transcription at 50°C followed by 2 min at 95°C. The following step was 40 cycles of 5 s each at at 95°C denaturation and 30 s at 60°C of primer annealing, extension, and relative fluorescence unit data collection. Data were analyzed with CFX Manager Software V3.0 (Bio[Rad, California, USA). *CCDC78* expression was compared to the expression of the reference genes. We evaluated the stability of potential reference genes in muscle tissue and PBLs: hypoxanthine phosphoribosyltransferase 1 (*HPRT1*, qHsaCID0016375), serpin peptidase inhibitor, clade C, member 1 (*SERPINC1*, qHsaCID0021147), glucuronidase, beta (*GUSB*, qHsaCIP0028142), connective tissue growth factor (*CTGF*, qHsaCED0002044), transferrin receptor (*TFRC*, qHsaCID0022106) and zinc finger protein 80 (*ZNF80*, qHsaCED0018708). *HPRT1*, *SERPINC1* and *ZNF80* were selected for data normalization, as they showed the most constant expression and stability. Relative quantification of gene expression was evaluated using the comparative threshold cycle value method 2^−ΔΔCt^.

### Western Blotting study

CCDC78 and RyR1 expression were evaluated by western blot (WB) analysis from muscle tissue obtained from the *CCDC78* mutated patient, four heterozygous *RYR1* mutated patients and three controls. Notably, two *RYR1* mutated patients harboured a mutation located in the CS of the protein (respectively c.130C>T (p.Arg44Cys) and c.1654C>T (p.Arg552Trp); the other two *RYR1* mutated patients carried a mutation located in channel and activation core of RyR1 (respectively c.11810C>T (p.Ser3937Leu) and c.11708G>A (p.Arg3903Gln)), according to the well know domains of the protein ^62^.

Muscle tissue (30-100 mg) was homogenized over ice using a Potter-type tissue homogenizer with Tissue Protein Extraction Reagent T-PER™ (Thermo Scientific, Rockford, USA) and Complete Mini Anti-protease Cocktail Tablets (Roche Applied Science, Laval, Canada) according to the manufacturer’s’ instructions. Total protein concentration was measured using Qubit^TM^ protein assay kit (Thermo Scientific, Rockford, USA). We prepared 50 μg protein for each sample through 5%, 12% and 4–15% Mini-PROTEAN® TGX™ precast gels (Bio[Rad, California, USA), and electrophoretically transferred to nitrocellulose membranes (Bio[Rad, California, USA). Membranes were blocked 1 h at room temperature (RT) in TBS containing 5% skimmed milk and 0.2% Tween 20, incubated overnight at 4°C with primary antibodies in 5% BSA and 0.2% Tween 20, and 1 h at RT with secondary antibodies in TBS containing 5% skimmed milk and 0.1% Tween 20. Antibodies were diluted as follows: 1:1700 rabbit anti-CCDC78 (AV53233, Sigma-Aldrich) 1:3300 rabbit anti-RyR1 ^63^, 1:20000 mouse anti-ß-actin (Santa Cruz Biotechnology, USA), 1:15000 mouse anti-GAPDH (39-8600, Thermo Scientific, Massachusetts, USA), 1:30000 (primary anti-CCDC78) and 1:80000 (primary anti-RyR1) peroxidase affinipure goat anti-rabbit IgG (111-035-045, Jackson ImmunoResearch Laboratories, Pennsylvania, USA) and 1:50000 peroxidase affinipure goat anti-mouse IgG (115-035-062, Jackson ImmunoResearch Laboratories, Pennsylvania, USA). Proteins were revealed by Clarity Max^TM^ Western ECL Substrate (Bio[Rad, California, USA) according to the manufacturer’s instructions and images acquisition from Gel Doc 2000 Imaging System (Bio[Rad, California, USA) were performed with Quantity One® 1-D analysis software. The relative amounts of CCDC78 and RyR1 were analyzed by densitometry using ImageJ software (https://rsb.info.nih.gov/ij). GAPDH was used to normalize the results. When necessary, the antibody was stripped with Restore Western Blot Stripping Buffer (Thermo Scientific). Data represent the mean of five and more than two independent replicates respectively for CCDC78 and RYR1 expression. Statistical analysis was performed by a two-tailed Student’s t-test; p-values of less than 0.05 and 0.01 were considered to be statistically significant and highly statistically significant, respectively.

### Co-immunoprecipitation assay and Western Blot assay

Fifty mg of muscle tissue from healthy controls and CCDC78 patient were lysed over ice using a Potter-type tissue homogenizer with IP Lysis/Wash Buffer (Thermo Scientific, Rockford, IL, USA) and Complete Mini Anti-protease Cocktail Tablets (Roche Applied Science, Laval, PQ, Canada) according to the manufacturer’s’ instructions. Lysates were incubated on ice for 5 minutes with periodic mixing and then cell debris were removed by centrifugation at 13,000×g for 10 minutes. Total protein concentration was measured using Qubit protein assay kit (Thermo Scientific). Co-Ip assay was performed using the Pierce Crosslink Magnetic IP/Co-IP Kit (Thermo Scientific, Rockford, IL, USA). Anti-CCDC78 (AV53233, Sigma-Aldrich) was diluted to the final concentration of 10µg/100µL. We used the crosslinking chemistry of disuccinimidyl suberate (DSS) to performe co-IP by coupling antibodies to the beads covalently. The lysate solution, diluted to 500µL with IP Lysis/Wash Buffer to a final concentration of 5 ng/ul of protein, was added to the tube containing crosslinked magnetic beads and incubated for 2,5 hours at 4°C on a rotator. Then we collected the beads and removed the unbound sample.

The immunoprecipitated protein was loaded in 5%, 12% and 4-15% gels. Gel was fixed (fixing solution: 50% methanol and 10% glacial acetic acid) for 1 hr with gentle agitation and then it was stained (staining solution: 0.1% Coomassie Brilliant Blue R-250, 50% methanol and 10% glacial acetic acid) for 20 min with gentle agitation. Afterwards the gel was destained (destaining solution: 40% methanol and 10% glacial acetic acid) and stored (storage solution: 5% glacial acetic acid) for mass spectrometry analysis.

For each immunoprecipitation assay, positive controls of protein expression at the whole lysate before co-IP and negative controls with nonspecific IgG (111-035-045, Jackson ImmunoResearch Laboratories, Pennsylvania, USA) were performed.

Co-IP elution (25-50 ul) were separated on 5%, 12% and 4-15% Mini-PROTEAN® TGX™ precast gels using methods described above. For protein blotting, the following primary antibodies were used: 1:1700 rabbit anti-CCDC78 (AV53233, Sigma-Aldrich), 1:3300 rabbit anti-RyR1 ^54^, 1:15000 mouse anti-GAPDH (39-8600, Thermo Scientific), 1:1000 mouse anti-ATP2A1 (VE121G9, Thermo Scientific), 1:3000 rabbit anti-ACTN2 (HPA008315, Sigma-Aldrich), 1:500 rabbit anti-TPM1 (HPA000261, Sigma-Aldrich), 1:1000 rabbit anti-CASQ1 (HPA026823, Sigma-Aldrich), 1:500 rabbit anti-TPM2 (11038-1-AP, Thermo Scientific), 1:1000 rabbit anti-MYH1 (ZRB1214, Sigma-Aldrich), 1:1000 mouse anti-ACTA1 (SAB4200602. Sigma-Aldrich), peroxidase affinipure goat anti-rabbit IgG (111-035-045, Jackson ImmunoResearch Laboratories) and peroxidase affinipure goat anti-mouse IgG (115-035-062, Jackson ImmunoResearch Laboratories).

Proteins were revealed by Clarity MaxTM Western ECL Substrate (Bio[Rad) according to the manufacturer’s instructions and images acquisition from Gel Doc 2000 Imaging System (Bio[Rad) were performed with Quantity One® 1-D analysis software. For each immunoprecipitation assay, positive controls of protein expression at the whole lysate before co-IP and negative controls with nonspecific IgG (111-035-045, Jackson ImmunoResearch Laboratories, Pennsylvania, USA) were performed. GAPDH was used as a negative control to demonstrate the specificity of co-IP.

### Mass spectroscopy study

The blue Coomassie stained 1D-gel was used for protein analysis. Seven bands of interest were selected and have been manually excised from the gel for the mass spectrometric analyses. Each band of interest underwent destaining and successive reduction, alkylation, and enzymatic digestion^64^ (see supplementary information). The final solution of the digested proteins was used for mass spectrometry experiments.

### nLC-nESI-HRMS/MS

The enzymatic digested solution obtained from gel band was analyzed by nano liquid chromatography (nLC) coupled to high resolution mass spectrometry (HRMS) equipped with a nanoelectrospray (nESI) interface (nLC-nESI-HRMS/MS)^65^. The instrument was composed by a nanoLC system EASY-nLC 1200 connected to a LTQ Orbitrap hybrid mass spectrometer (Thermo Scientific, Bremen, Germany). Excalibur softwares were used for instruments control and data acquisition (version 4.2 for EASY-nLC and version 2.0.7 for LTQ Orbitrap).The nESI interface parameters were: nESI spray potential 1.7 kV, capillary and tube lens voltages were 42 and 120 V, respectively. MS data acquisition was done in DDA (data dependent acquisition) mode, performing a HRMS (60000 nominal resolution, at m/z 400) full scan from 350 to 2000 m/z in the Orbitrap analyzer, using a 1 x 106 target value. MS/MS spectra were recorded in the linear quadrupole ion trap analyzer: precursor ions were selected from the 7 most intense signals in the HRMS full scan spectrum above 600 a.u. threshold and with an isolation window of 2.2 Da. Normalized collision energy of 35% and 20 ms activation time were used. Singly charged or no charge state assigned precursor ions did not trigger MS/MS experiments. Dynamic exclusion of already selected precursor masses was applied, with an exclusion time of 30 s and an exclusion window of 20 ppm (repeat count 2, repeat duration 15 s). The chromatographic column was an AcclaimÒ PepMap 100 C18, 3 µm, 100 Å, 75 µm x 150 mm (Dionex Thermo), operating at 0.3 μL/min flow rate. Solvent A was 100 % water and solvent B was 80% acetonitrile/20% water, both containing 0.1% formic acid; solvents were of LC-MS grade from Sigma (Sigma Italy, Merck). A 1 µL volume was injected into the nLC-nESI HRMS/MS system. Elution was done by gradient starting from 2% B for 5 min, to 40% B in 340 min, to 90% B in 5 min and then returned to initial conditions.

### Filtering and selecting interactors candidates for validation

The acquired data were analyzed with Mascot 2.4 search engine (Matrix Science Ltd., London, UK) against a human database created from NCBI. Searches were performed allowing: (i) trypsin as enzyme, (ii) up to two missed cleavage sites, (iii) 10 ppm of tolerance for the monoisotopic precursor ion and 0.5 mass unit for monoisotopic fragment ions, (iv) carbamidomethylation of cysteine and oxidation of methionine as variable modifications. A target-decoy search was used: a false discovery rate (FDR) of 1% was imposed and the criterion used to accept protein identification included probabilistic score sorted by the software. We identifed a total of 478 known database entries, after we eliminated proteins contaminants such as the keratin family and immunogubulin to obtain 367 total interactors with number of unique peptides per protein ≥1.

### Three-dimensional (3D) modelling

Three-dimensional (3D) modelling was performed and generated through SwissModel (https://swissmodel.expasy.org/) in order to explain the possible effect of the mutation on CCDC78 protein structure. The analysis used the human template A2IDD5.1.A (GMQE=0.66) for the 48 kDa isoform; as for the 52 kDa isoform, the software used the near-atomic resolution structure of the almost identical (70%) S9XHM5.1.A template of Camelus ferus (GMQE=0.69). Moreover, using DAS method (https://tmdas.bioinfo.se/)^10^, we analyzed the possible presence of transmembrane domain in CCDC78 protein. We obtained curves by pairwise comparison of the proteins in the test set in “each against the rest” fashion. There are two cutoffs indicated on the generated plots: a “strict” one at 2.2 DAS score, and a “loose” one at 1.7. The hit at 2.2 is informative in terms of the number of matching segments, while a hit at 1.7 gives the actual location of the transmembrane segment. The segments reported in the feature table (FT) records of the SwissProt database are marked at 1.0 DAS score (”FT lines”). Project HOPE, a web server that investigates the structural consequences of variants by cooperating with UniProt and DAS prediction algorithms^66^, was used to study p.Glu404Lys mutation in NM_001378030.1 isoform.

## Data Availability

All data produced in the present study are available upon reasonable request to the authors

## Acknowledgements

This work has been supported by the “InGene 2.0” grant (“Bando Ricerca Salute 2018,” Decree n. 15397, 26–09-2018, Tuscany Region) (A.M., F.M.S.), AFM-Telethon grant n. 24835 (D.R.), PNRR CN3, spoke 1 (V.S.), Ricerca Corrente 2023 - Ministry of Health (F.M.S.). Dr. Diego Lopergolo and Prof. Alessandro Malandrini are members of the ERN Euro-NMD.

## References

1. Cassandrini, D. et al. Congenital myopathies: Clinical phenotypes and new diagnostic tools. Ital. J. Pediatr. 43, (2017).

2. North, K. N. et al. Approach to the diagnosis of congenital myopathies. Neuromuscul. Disord. 24, (2014).

3. Papadimas, G. K., Xirou, S., Kararizou, E. & Papadopoulos, C. Update on congenital myopathies in adulthood. International Journal of Molecular Sciences vol. 21 (2020).

4. Volk, A. E. & Kubisch, C. The rapid evolution of molecular genetic diagnostics in neuromuscular diseases. Current Opinion in Neurology vol. 30 (2017).

5. Daniels, R. J. et al. Sequence, structure and pathology of the fully annotated terminal 2 Mb of the short arm of human chromosome 16. Hum. Mol. Genet. 10, (2001).

6. Tang, T. K. Centriole biogenesis in multiciliated cells. Nat. Cell Biol. 15, 1400–1402 (2013).

7. Kim, S. K. et al. A role for Cep70 in centriole amplification in multiciliated cells. Dev. Biol. 471, 10–17 (2021).

8. Majczenko, K. et al. Dominant mutation of CCDC78 in a unique congenital myopathy with prominent internal nuclei and atypical cores. Am. J. Hum. Genet. 91, (2012).

9. Gonorazky, H. D. et al. Expanding the Boundaries of RNA Sequencing as a Diagnostic Tool for Rare Mendelian Disease. Am. J. Hum. Genet. 104, (2019).

10. Cserzo, M., Wallin, E., Simon, I., von Heijne, G. & Elofsson, A. Prediction of transmembrane alpha-helices in prokaryotic membrane proteins: the dense alignment surface method. Protein Eng. Des. Sel. 10, 673–676 (1997).

11. Vasli, N. et al. Recessive mutations in the kinase ZAK cause a congenital myopathy with fibre type disproportion. Brain 140, (2017).

12. Rossi, D. et al. The Sarcoplasmic Reticulum of Skeletal Muscle Cells: A Labyrinth of Membrane Contact Sites. Biomolecules vol. 12 (2022).

13. Dulhunty, A. F. Excitation-contraction coupling from the 1950s into the new millennium. Clinical and Experimental Pharmacology and Physiology vol. 33 (2006).

14. Kawasaki, T. & Kasai, M. Regulation of calcium channel in sarcoplasmic reticulum by calsequestrin. Biochem. Biophys. Res. Commun. 199, (1994).

15. Manno, C. et al. Calsequestrin depolymerizes when calcium is depleted in the sarcoplasmic reticulum of working muscle. Proc. Natl. Acad. Sci. U. S. A. 114, (2017).

16. Gokhin, D. S. et al. Tropomodulin isoforms regulate thin filament pointed-end capping and skeletal muscle physiology. J. Cell Biol. 189, (2010).

17. Perni, S. & Beam, K. G. Voltage-induced Ca2+ release is supported by junctophilins 1, 2 and 3, and not by junctophilin 4. J. Gen. Physiol. 154, (2022).

18. Rossi, D. et al. Molecular determinants of homo- And heteromeric interactions of Junctophilin-1 at triads in adult skeletal muscle fibers. Proc. Natl. Acad. Sci. U. S. A. 116, (2019).

19. Sangkuhl, K., Dirksen, R. T., Alvarellos, M. L., Altman, R. B. & Klein, T. E. PharmGKB summary: Very important pharmacogene information for CACNA1S. Pharmacogenet. Genomics 30, (2020).

20. Amoasii, L. et al. Myotubularin and PtdIns3 *P* remodel the sarcoplasmic reticulum in muscle *in vivo*. J. Cell Sci. (2013) doi:10.1242/jcs.118505.

21. Srikanth, S. et al. Junctate is a Ca ^2^ ^+^ -sensing structural component of Orai1 and stromal interaction molecule 1 (STIM1). Proc. Natl. Acad. Sci. 109, 8682–8687 (2012).

22. Srikanth, S. et al. A novel EF-hand protein, CRACR2A, is a cytosolic Ca2+ sensor that stabilizes CRAC channels in T cells. Nat. Cell Biol. 12, (2010).

23. Shen, X. et al. Triadins modulate intracellular Ca2+ homeostasis but are not essential for excitation-contraction coupling in skeletal muscle. J. Biol. Chem. 282, (2007).

24. Zhang, L., Salgado-Somoza, A., Vausort, M., Leszek, P. & Devaux, Y. A heart-enriched antisense long non-coding RNA regulates the balance between cardiac and skeletal muscle triadin. Biochim. Biophys. Acta - Mol. Cell Res. 1865, (2018).

25. Yan, Z. et al. Structure of the rabbit ryanodine receptor RyR1 at near-atomic resolution. Nature 517, (2015).

26. Dulhunty, A. F., Beard, N. A. & Casarotto, M. G. Recent advances in understanding the ryanodine receptor calcium release channels and their role in calcium signalling [version 1; peer review: 4 approved]. F1000Research vol. 7 (2018).

27. Lamboley, C. R., Murphy, R. M., Mckenna, M. J. & Lamb, G. D. Sarcoplasmic reticulum Ca2+ uptake and leak properties, and SERCA isoform expression, in type I and type II fibres of human skeletal muscle. J. Physiol. 592, (2014).

28. Lamboley, C. R., Murphy, R. M., Mckenna, M. J. & Lamb, G. D. Endogenous and maximal sarcoplasmic reticulum calcium content and calsequestrin expression in type I and type II human skeletal muscle fibres. J. Physiol. 591, (2013).

29. Laing, N. G. et al. Mutations and polymorphisms of the skeletal muscle α-actin gene (ACTA1). Human Mutation vol. 30 (2009).

30. Monnier, N. et al. An autosomal dominant congenital myopathy with cores and rods is associated with a neomutation in the RYR1 gene encoding the skeletal muscle ryanodine receptor. Hum. Mol. Genet. 9, (2000).

31. Von Der Hagen, M., et al. Novel RYR1 missense mutation causes core rod myopathy. European Journal of Neurology vol. 15 (2008).

32. Hernandez-Lain, A. et al. De novo RYR1 heterozygous mutation (I4898T) causing lethal core-rod myopathy in twins. Eur. J. Med. Genet. 54, (2011).

33. Scacheri, P. C. et al. A novel ryanodine receptor gene mutation causing both cores and rods in congenital myopathy. Neurology 55, (2000).

34. Sparrow, J. C. et al. Muscle disease caused by mutations in the skeletal muscle alpha-actin gene (ACTA1). Neuromuscular Disorders vol. 13 (2003).

35. Beard, N. A., Laver, D. R. & Dulhunty, A. F. Calsequestrin and the calcium release channel of skeletal and cardiac muscle. Prog. Biophys. Mol. Biol. 85, 33–69 (2004).

36. Gautel, M. & Djinović-Carugo, K. The sarcomeric cytoskeleton: from molecules to motion. J. Exp. Biol. 219, 135–45 (2016).

37. Chopra, A. et al. Force Generation via β-Cardiac Myosin, Titin, and α-Actinin Drives Cardiac Sarcomere Assembly from Cell-Matrix Adhesions. Dev. Cell 44, 87–96.e5 (2018).

38. Sanger, J. W., Wang, J., Holloway, B., Du, A. & Sanger, J. M. Myofibrillogenesis in skeletal muscle cells in zebrafish. Cell Motil. Cytoskeleton 66, 556–66 (2009).

39. Wang, C., Yue, F. & Kuang, S. Muscle Histology Characterization Using H&amp;E Staining and Muscle Fiber Type Classification Using Immunofluorescence Staining. BIO-PROTOCOL 7, (2017).

40. Valberg, S. J., Henry, M. L., Perumbakkam, S., Gardner, K. L. & Finno, C. J. An E321G *MYH1* mutation is strongly associated with nonexertional rhabdomyolysis in Quarter Horses. J. Vet. Intern. Med. 32, 1718–1725 (2018).

41. Alsaif, H. S. et al. MYH1 is a candidate gene for recurrent rhabdomyolysis in humans. Am. J. Med. Genet. A 185, 2131–2135 (2021).

42. Eckhardt, J. et al. Quantitative proteomic analysis of skeletal muscles from wild-type and transgenic mice carrying recessive Ryr1 mutations linked to congenital myopathies. Elife 12, (2023).

43. Wang, X., Kong, C., Liu, P., Geng, W. & Tang, H. Identification of Potential Biomarkers for Ryanodine Receptor 1 (RYR1) Mutation-Associated Myopathies Using Bioinformatics Approach. Dis. Markers 2022, 8787782 (2022).

44. Wang, E. T. et al. Alternative isoform regulation in human tissue transcriptomes. Nature 456, (2008).

45. Zhang, Z., Miteva, M. A., Wang, L. & Alexov, E. Analyzing effects of naturally occurring missense mutations. Computational and Mathematical Methods in Medicine vol. 2012 (2012).

46. Boulling, A. et al. Functional analysis of pancreatitis-associated missense mutations in the pancreatic secretory trypsin inhibitor (SPINK1) gene. Eur. J. Hum. Genet. 15, (2007).

47. Klos Dehring, D. A., et al. Deuterosome-Mediated Centriole Biogenesis. Dev. Cell 27, 103–112 (2013).

48. Ng, D. C. H., Ho, U. Y. & Grounds, M. D. Cilia, Centrosomes and Skeletal Muscle. Int. J. Mol. Sci. 22, 9605 (2021).

49. Palla, A. R. et al. Primary cilia on muscle stem cells are critical to maintain regenerative capacity and are lost during aging. Nat. Commun. 13, 1439 (2022).

50. Marg, A., Haase, H., Neumann, T., Kouno, M. & Morano, I. AHNAK1 and AHNAK2 are costameric proteins: AHNAK1 affects transverse skeletal muscle fiber stiffness. Biochem. Biophys. Res. Commun. 401, 143–148 (2010).

51. Liu, S., Li, S., Zhu, H., Cheng, S. & Zheng, Q. Y. A mutation in the cdh23 gene causes age-related hearing loss in Cdh23nmf308/nmf308 mice. Gene 499, 309–317 (2012).

52. Dafinger, C. et al. Mutations in KIF7 link Joubert syndrome with Sonic Hedgehog signaling and microtubule dynamics. J. Clin. Invest. 121, (2011).

53. Tomelleri, G. et al. SERCA1 and calsequestrin storage myopathy: A new surplus protein myopathy. Brain 129, (2006).

54. Rossi, D. et al. A Mutation in the CASQ1 Gene Causes a Vacuolar Myopathy with Accumulation of Sarcoplasmic Reticulum Protein Aggregates. Hum. Mutat. 35, (2014).

55. Genovese, I. et al. Sorcin is an early marker of neurodegeneration, Ca2+ dysregulation and endoplasmic reticulum stress associated to neurodegenerative diseases. Cell Death Dis. 11, (2020).

56. Vorobjev, I. A., Uzbekov, R. E., Komarova, Y. A. & Alieva, I. B. γ-Tubulin distribution in interphase and mitotic cells upon stabilization and depolymerization of microtubules. Membr. Cell Biol. 14, (2000).

57. Abràmoff, M. D., Magalhães, P. J. & Ram, S. J. Image processing with imageJ. Biophotonics International vol. 11 (2004).

58. Bøyum, A. [9] Separation of Lymphocytes, Granulocytes, and Monocytes from Human Blood Using Iodinated Density Gradient Media. Methods Enzymol. 108, (1984).

59. Mortazavi, A., Williams, B. A., McCue, K., Schaeffer, L. & Wold, B. Mapping and quantifying mammalian transcriptomes by RNA-Seq. Nat. Methods 5, (2008).

60. Anders, S. & Huber, W. Differential expression analysis for sequence count data. Genome Biol. 11, (2010).

61. Trapnell, C. et al. Transcript assembly and quantification by RNA-Seq reveals unannotated transcripts and isoform switching during cell differentiation. Nat. Biotechnol. 28, (2010).

62. des Georges, A. et al. Structural Basis for Gating and Activation of RyR1. Cell 167, (2016).

63. Rossi, D. et al. Distinct regions of triadin are required for targeting and retention at the junctional domain of the sarcoplasmic reticulum. Biochem. J. 458, (2014).

64. Rappsilber, J., Mann, M. & Ishihama, Y. Protocol for micro-purification, enrichment, pre-fractionation and storage of peptides for proteomics using StageTips. Nat. Protoc. 2, (2007).

65. Dani, F. R. & Pieraccini, G. Proteomics of arthropod soluble olfactory proteins. in Methods in Enzymology vol. 642 (2020).

66. Venselaar, H., te Beek, T. A., Kuipers, R. K., Hekkelman, M. L. & Vriend, G. Protein structure analysis of mutations causing inheritable diseases. An e-Science approach with life scientist friendly interfaces. BMC Bioinformatics 11, 548 (2010).

