## Supplementary information for "CCDC78: unveiling the function of a novel gene associated to hereditary myopathy"

**Results**

**RNA-seq analyses**

Although statistically significant only according to p-values and not adjusted p-values, among the subsequent 647 processes we found: positive regulation of skeletal muscle fiber development (488th position), positive regulation of muscle cell differentiation (517th), muscle cell development (541th), striated muscle cell differentiation (581th), skeletal muscle fiber development (638th), intrinsic apoptotic signaling pathway in response to endoplasmic reticulum stress (646th), regulation of muscle contraction (651th), negative regulation of muscle hypertrophy (833th), positive regulation of fast-twitch skeletal muscle fiber contraction (850th), establishment of protein localization to endoplasmic reticulum (926th), negative regulation of muscle tissue development (942th), negative regulation of skeletal muscle hypertrophy (953th), endoplasmic reticulum calcium ion homeostasis (984th), muscle filament sliding (998th), skeletal muscle tissue regeneration (1013th).

**Mass spectroscopy study**

**Table S1. Identified possible CCDC78 partners by nLC-nESI-HRMS/MS analysis.**

| **Band n.** | **Accession** | **Score** | **Mass** | **Num. of significant matches** | **Num. of significant sequences** | **emPAI** | **Description** |
| --- | --- | --- | --- | --- | --- | --- | --- |
| 1 | MYH1_HUMAN | 4021 | 223006 | 200 | 86 | 3,27 | Myosin-1 GN=MYH1 |
| 2 | AT2A1_HUMAN | 286 | 110182 | 19 | 13 | 0,46 | Sarcoplasmic/endoplasmic reticulum calcium ATPase 1 GN=ATP2A1 |
| 3 | ACTN2_HUMAN | 2086 | 103788 | 98 | 44 | 4,03 | Alpha-actinin-2 GN=ACTN2 |
| 4 | CASQ1_HUMAN | 168 | 45132 | 8 | 5 | 0,42 | Calsequestrin-1 GN=CASQ1 |
| 5 | ACTS_HUMAN | 2427 | 42024 | 133 | 22 | 9,43 | Actin, alpha skeletal muscle GN=ACTA1 |
| 6 | TPM2_HUMAN | 1211 | 32831 | 65 | 23 | 10,1 | Tropomyosin beta chain GN=TPM2 |
| 7 | TPM1_HUMAN | 1327 | 32689 | 78 | 29 | 21,22 | Tropomyosin alpha-1 chain GN=TPM1 |

**Legend**: emPAI (exponentially modified protein abundance index); Score (overall Protein Score reflects the combined scores of all observed mass spectra that can be matched to amino acid sequences within that protein. A higher score indicates a more confident match).

**Materials and methods**

**Morphological analysis of muscle biopsies and HeLa cells**

For ultrastructural studies, small samples were fixed in 2.5% glutaraldehyde, pH 7.4, postfixed in 2% osmium tetroxide for 2 hours, dehydrated and embedded in epoxy resin. At least 3 blocks from each sample were studied, including longitudinal and transverse-oriented samples. Semi-thin sections were stained by toluidine blue and examined with a Zeiss Axiomat light microscope to select pathological areas. Ultrathin sections were stained with uranyl acetate and lead citrate. The grids were observed using a Zeiss EM 109 electron microscope (80 kV; Karl Zeiss, Berlin, Germany).

**DNA analysis**

WES was performed by the NovaSeq6000 (Illumina) on DNA from peripheral blood samples. Mutational analysis was carried on using GATK version v.4.0. External datasets, such as 1000 genomes, ExAC and GnomAD, were used to define novel variants, not previously identified. We performed a prioritization of the variants selecting frameshift, splice, stopgain or stoploss mutations, missense variants predicted to be damaging by CADD-phred prediction tools and variants with minor allele frequency (MAF) < 0.01. Single nucleotide variants were confirmed by Sanger sequencing. The American College of Medical Genetics and Genomics (ACMG) criteria were used to classify detected variants. Putatively deleterious variants were validated by PCR-based standard capillary Sanger sequencing.

The exons and adjacent intron regions of *CCDC78* gene were amplified by PCR using primers specific for *CCDC78* gene. The PCR were sequenced in both forward and reverse directions by automated sequencer ABI 3500 (Applied Biosystems). The results were analyzed using Chromas version 2.33 software and compared with reference sequence NG_032932.1. Mutation was numbered according to the published cDNA sequence NM_001378030.1 for the longer transcript and according to NM_001031737.3 for the shorter transcript.

**Primary fibroblast cultures**

Primary cultures of fibroblasts, obtained from healthy control skin biopsies were grown in standard condition with 1 ml of Dulbecco's modified Eagle's medium (Merck KGaA, Darmstadt, Germany) supplemented with 10% fetal calf serum (Merck KGaA), 1% l-glutamine, and 1% streptomycin-penicillin (100 IU/ml and 100 µg/ml, respectively; Merck KGaA). Flasks were maintained at 37°C in a humidified atmosphere containing 5% CO_2_. Analysis of subconfluent fibroblasts cultures was carried out after 15 replications. At this time, cells were treated with 2 cc of trypsin and harvested in complete medium.

**HeLa cells**

HeLa cells were grown in a 5% CO_2_ atmosphere in Dulbecco’s modified Eagle’s medium high glucose (DMEM; Euroclone), supplemented with 10% fetal bovine serum (Gibco), 100 U/ml penicillin and 100 mg/ml streptomycin.

**Transcripts analysis**

PCR amplifications were performed with primers surrounding the mutated region (TabS2).

**Tab. S2. List of primers used to amplify exons 10-14 region of *CCDC78* gene**

| CCDC78-10/11RNAFW3 | CAGGCAGTGGAGCACGCAGAT |
| --- | --- |
| CCDC78-11/12RNAFW | GGAGGACCAGCACGGCGG |
| CCDC78-12/13RNAFW | CATCAGAGCCACAGGGCCTG |
| CCDC78-14RV | CGTGCTTGTACCTGCCCAGGT |
| CCDC78-14RV2 | TCAGGATTTCGTGCTTGT |

**Mass spectroscopy study**

The blue Coomassie stained 1D-gel was used for protein analysis. Seven bands of interest were selected and have been manually excised from the gel using a clean scalpel blade for the following mass spectrometric analyses. Each band of interest was transferred into a 1.5 mL low-binding Eppendorf tube for destaining and successive reduction, alkylation, and enzymatic digestion.

Destaining was performed by adding 120 μL of ACN to the gel band, sufficient to cover the gel piece, and left for ten minutes; then the ACN was discarded and 100 μL 0.1 M NH4HCO3 were added. After 5 minutes NH4HCO3 solution was removed. These two steps were repeated two times or more until the supernatant was no more colored. A final 100 μL volume of ACN was added and then discarded and the residual ACN was evaporated into a centrifugal evaporator concentrator (Jouan, Thermo Fisher). To the gel band, 100 μL of 10 mM DTT in 0.1 M NH4HCO3 were added and the Eppendorf tube was left at 56 °C for 45 min in an orbital shaker. The sample was taken to room temperature and the solution removed; 130 μL of ACN were added, vortexed, and then immediately discarded. Alkylation was done by adding 100 μL of 55 mM iodoacetamide in 0.1 M NH4HCO3; the solution was shaken for 30 min at room temperature, protected from the light. The iodoacetamide solution was removed and then 100 μL of ACN were added, then discarded; 100 μL of 0.1 M NH4HCO3 were added and, after shaking for 10 minutes, discarded. These steps were repeated two times and then 100 μL of 100% ACN were added to each tube. After shaking for 5 min, the ACN was removed and left to evaporate the residual ACN for 20 min. The enzymatic digestion was performed by adding 50 μL of 50 mM NH4HCO3 and 1 μL of 0.4 μg/μL LysC solution in ultrapure water. The sample was left for 3 h at 37 °C under shacking. Then 1 μL of 0.5 μg/μL trypsin solution in 50 mM acetic acid was added and left to incubate overnight at 37 °C under shacking. The reaction was stopped by adding 1 μL of 10% TFA and the pH was measured to ensure that it was around 3; more 10% TFA was added if necessary. The sample was centrifuged and the supernatant was used for C18 STAGE tip purification, following the procedure described by Rappsilber et al.

The C18 STAGE tip was eluted directly in a polypropylene conical insert with 50 μL of 0.5% acetic acid in H2O:ACN 20:80 (v:v). The solution was concentrated under vacuum in a centrifugal evaporator to approximately 10 μL and then the final volume was brought to 20 μL with 0.5% acetic acid. The final solution of the digested proteins was used for mass spectrometry experiments.
